# Longitudinal Multi-omics Reveals Persistent Gut Microbial and Metabolic Dysbiosis Following Anti-Tuberculosis Therapy

**DOI:** 10.64898/2026.10.05.26364754

**Authors:** Archana Pant, Sumeet Kumar Yadav, Jayalakshmi Srinivasan, Sonu Kumar Gupta, Bhanu Pratap Gurjar, Pradipta Jana, Daizee Talukdar, Senbagavalli Prakash Babu, Sonali Sarkar, Yashwant Kumar, Debasisa Mohanty, Bhabatosh Das, G. Aneeshkumar Arimbasseri

## Abstract

Pulmonary tuberculosis (TB) is associated with systemic alterations extending beyond the lungs, yet the longitudinal impact of anti-tuberculosis therapy (ATT) on gut ecosystem recovery remains poorly understood. Here, we performed a longitudinal multi-omics analysis of gut microbial and metabolic dynamics in pulmonary TB patients (n=8) followed at baseline, 1, 6, and 8 months of ATT, compared with healthy household contacts (HHCs; n=8). Shotgun metagenomics, functional profiling, untargeted fecal metabolomics, resistome, plasmidome, virome, and microbiome-metabolome integration analyses were performed to characterize disease-associated alterations and treatment-associated recovery. Active, treatment-naive TB was characterized by reduced microbial diversity, altered community structure, depletion of beneficial commensals, and disruption of microbial functional pathways and metabolic profiles. During ATT, microbial diversity and functional/metabolic features progressively recovered; however, post-treatment profiles remained distinct from HHCs, indicating incomplete ecological restoration after microbiological cure. Microbiome-metabolome analysis revealed associations between altered bacterial taxa and metabolites involved in amino acid metabolism, bile acid transformation, oxidative stress, and host-microbial metabolic interactions. In contrast, resistome and mobile genetic element profiles demonstrated greater stability throughout treatment. HHCs exposed to index TB cases maintained relative gut ecosystem stability over time. Bacterial-AMR correlation analysis identified taxon-specific resistance associations among gut bacteria. Together, these findings provide a longitudinal systems-level view of gut ecosystem remodeling during TB and highlight that restoration of microbial homeostasis extends beyond pathogen clearance.

## INTRODUCTION

Pulmonary tuberculosis, caused by *Mycobacterium tuberculosis* (Mtb), remains a dominant global infectious threat, necessitating intensive, long-term multi-drug regimens to ensure microbiological clearance and prevent the emergence of systemic drug resistance. Standard frontline antituberculosis therapy (ATT), typically comprising a combination of rifampicin, isoniazid, ethambutol, and pyrazinamide, must be sustained continuously for six months or longer^1^. While this aggressive drug intervention is clinically optimized to eliminate slow-growing mycobacterial populations in the lung, its prolonged duration exerts profound, continuous xenobiotic pressure on the host’s distal gastrointestinal tract. The human intestinal lumen harbors a dense, complex consortium of obligate anaerobes that maintain metabolic homeostasis and mucosal barrier immunity. Under the collateral exposure of broad-spectrum frontline ATT agents, this homeostatic ecosystem inevitably undergoes severe structural and functional reconfigurations^2,3,4,5,6^.

Emerging paradigms in microbial ecology indicate that the true public health risk of antibiotic-mediated dysbiosis extends far beyond the transient depletion of beneficial taxa^7^. Rather, the intestinal tract acts as a highly dynamic resistome, a foundational reservoir of antibiotic resistance genes (ARGs) capable of adapting rapidly to xenobiotic stress^8^. During long-term multi-drug exposure, the absolute quantitative load of this resistome can surge, trapping the host in a persistent, narrow state dominated by multi-drug-resistant determinants. Crucially, the tempo and permanence of this resistome inflation are not dictated solely by vertical bacterial inheritance. Instead, horizontal gene transfer (HGT), orchestrated by a diverse suite of mobile genetic elements (MGEs) such as plasmids, viruses, transposons, and integrons, serves as the primary evolutionary engine driving the rapid dissemination of resistance across disparate phylogenetic boundaries^9,10^.

Crucially, the ecological dynamics of this resistome inflation do not occur in isolation. In endemic settings, active tuberculosis transmission necessitates prolonged, close contact co-habitation within a shared environment. Household contacts might share not only a physical space and dietary habits with active patients but also a continuous baseline exchange of ambient microflora^11,12^. Tracking these longitudinal healthy controls over an identical six-month monitoring window provides an invaluable epidemiological control group. This dual-cohort framework decouples the independent structural impacts of shared household pathogen exposure, environmental clustering, and natural temporal drift from the direct, systemic selective pressures exerted by multi-drug therapy. Characterizing how the background resistome and mobilome fluctuate in healthy co-habiting contacts over time is essential for defining true homeostatic baselines, ensuring that longitudinal reconfigurations identified in the patient cohorts are definitively recognized as a direct evolutionary consequence of ATT rather than shared household environmental confounding.

While standard antibiotic perturbations are known to reduce overall intestinal bacterial diversity, their precise long-term impact on AMR-MGE diversity, absolute abundance trajectories, and subsequent mobilome-resistome coupling remains fundamentally unresolved in the context of chronic human pathogenesis. Specifically, how the intestinal resistome and mobilome interact under the specific selective pressures of prolonged anti-mycobacterial agents, and whether these communities can ever revert to a homeostatic baseline post-therapy, remains a critical knowledge gap. Understanding these cooperative dynamics is of paramount clinical importance; the collateral enrichment of highly mobile resistance networks within commensal populations could inadvertently cultivate an absolute breeding ground for opportunistic, multi-drug resistant pathobionts, ultimately complicating post-treatment secondary infections and long-term patient recovery.

To address these critical questions, this study systematically tracks the longitudinal multi-omics landscape of the human gut ecosystem across six distinct clinical cohorts, encompassing baseline healthy, household contacts, active treatment-naive tuberculosis patients, and individuals at intermediate and post-ATT monitoring phases. Leveraging deep metagenomic sequencing alongside untargeted stool metabolomics, we comprehensively map the parallel evolution of gut taxonomy, absolute ARG burdens, and MGE diversity tracks. Furthermore, through network-based interactomics, we chart the explicit physical and functional linkages bridging specific bacterial lineages, mobile genetic elements, and metabolic stress phenotypes. Our findings reveal that Mtb infection reconfigures the intestinal ecosystem prior to drug exposure, and that subsequent ATT drives a coordinated, long-term consolidation of the gut mobilome and resistome. This structural shift locks the intestinal lumen into a narrow, pathobiont-fuelled resistance state that persists long after active therapy concludes, redefining our understanding of the collateral evolutionary costs of standard tuberculosis management.

## MATERIALS AND METHODS

### Subject enrollment and sample collection

All study subjects were recruited at Jawaharlal Institute of Postgraduate Medical Education and Research (JIPMER), Pondicherry, India, following written informed consent. The study protocol and recruitment criteria were approved by the institutional ethics committees of BRIC-National Institute of Immunology (NII), Delhi and JIPMER. Patients with pulmonary tuberculosis (TB) were prospectively recruited based on clinical symptoms and confirmed by sputum-smear microscopy. Drug-sensitive TB patients were initiated on the WHO-recommended Directly Observed Treatment, Short-course (DOTS) regimen under the National TB Elimination Programme (NTEP), comprising isoniazid, rifampicin, ethambutol, and pyrazinamide. Stool samples were collected longitudinally from TB patients before anti-TB treatment (M0, *n*=7), after one month of treatment (M1, *n*=8), at treatment completion (M6, *n*=8), and two months post-treatment (M8, *n*=7) (One sample missing at M0 and M8). One healthy household close contact of each patient was enrolled as a control (*n*=8), and stool samples were collected at M0 and M6 (both *n*=8), corresponding to the baseline and treatment completion time points of the TB cohort. Samples were transported to BRIC-NII, Delhi, and stored at -80°C until further processing. Demographic and clinical information, including vital signs, dietary intake, and medical history, was recorded at each sampling visit. Peripheral blood was also collected from all participants at each study time point for complete blood count (CBC) analysis. To study the complete blood count (CBC) pattern of all participants, the blood samples were also collected at all study time-points. The raw values of 14 CBC parameters were log_10_-transformed for all the 6 groups. Each parameter was analysed separately for all the 6 groups using paired-t-test (H_M0 vs H_M6), unpaired-t-test (H_M0 vs P_M0), and mixed-effects analysis through One-way Anova (P_M0, P_M1, P_M6, P_M8). The analysis was done using GraphPad Prism 8.

### Isolation of Metagenomic DNA from stool samples and sequencing

Approximately, 500 mg of frozen stool sample was taken to extract metagenomic DNA (mgDNA) using THSTI DNA extraction method^13^ (a combinatorial method consisting of enzymatic, chemical and mechanical lysis for proper lysis of microbial content). Quality and quantity of the isolated mgDNA(s) were monitored by resolving the sample in 0.8% agarose gel. The 260/230 and 260/280 ratios were used to check the mgDNA purity. The extracted DNA samples were used for metagenome sequencing by Neuberg Diagnostics (India). Briefly, 250 ng of total DNA per sample was used as an input material for sequencing library construction using a KAPA Hyper Plus Kit (Roche #07962428001). After quality assessment, the library preparations were sequenced on an Illumina NovaSeq 6000 platform, and paired-end raw reads were generated with a read length of 150 bps.

### Sequencing data processing and inferring gut microbiota composition

Raw sequencing reads were quality-checked using FastQC (v0.12.1) and trimmed using fastp (v0.23.4). The sequencing data generated a total of 1874.912304 M reads from 46 samples, with a minimum of 19.30463 M and a maximum 60.612274 M reads after quality filtering. The average number of reads generated were found to be 40.75896313 M (Table S1). The sample reads with human contamination were identified and removed if they were ≥1% by mapping against the human genome (hg19) with Bowtie2 (v2.5.3), Samtools (v1.23) and Bedtools (v2.29.1). Quality-filtered metagenomes were taxonomically profiled using Kraken2 (v2.1.3) with default parameters and assembled with MegaHit tool (v1.2.9). Kraken reports were used for abundance and diversity analysis and LEfSe identified exploratory microbial features. The taxa below 0.01% abundance were removed from analysis. Functional profiling of the metagenomic data was performed using HUMAnN (v3.9) on complete data and exploratory microbial pathways were identified by LEfSe analysis using the Microeco R package.

### Identification and analysis of ARGs and MGEs in metagenome data

For antimicrobial resistance (AMR) profiling, reads were mapped directly to core CARD database (v4.0.1) using RGI-bwt (v6.0.3). AMR genes with ≥ 80% coverage, a minimum of 10 reads mapped and present in atleast 5% of samples were retained for further analysis. For mobile genetic elements (MGEs), i.e. plasmids and viral identification and analysis, MegaHit assembled contigs were screened using geNomad (v1.8.0) with the *–enable-score-calibration* option. For plasmidome analysis, the plasmid sequences identified using geNomad (v1.8.0) were classified as follows. Circular plasmids were sequences meeting the criteria of length >1 kb, FDR <0.05 (estimated using *–enable-score-calibration)*, and the presence of direct terminal repeats (DTRs). Plasmid fragments were sequences with length >1 kb, FDR <0.05 and no DTRs. Circular plasmids and plasmid fragments were then dereplicated separately and clustered into PTUs using the CheckV scripts *anicalc.py* and *aniclust.py* with strict (*–min_ani 95 –min_tcov 85 –min_qcov 85*) and loose (*–min_ani 90 –min_tcov 85 –min_qcov 0)* parameters, respectively. Both plasmid sets were then combined using the loose settings, filtering fragment PTUs that clustered together with circular PTUs. For virus prediction, viral sequences meeting the criteria of length >1 kb, FDR <0.05 (estimated using *–enable-score-calibration)* were considered. CheckV (v1.1.0) was used to further trim predicted viral sequences to remove potential host regions, and assess genome quality of resulting sequences using the end-to-end pipeline and select contigs that met ≥50% completeness criteria. Medium and high quality (total 619) sequences were considered for downstream analysis. Dereplication was performed using the MIUVIG recommended settings (95% ANI; 85% AF (aligned fraction)), using the CheckV scripts *anicalc.py* and *aniclust.py* with the ‘–min_ani 95 –min_tcov 85 –min_qcov’ parameters. The longest sequence in each cluster was selected as the vOTU representative^14,15^. Dereplication yielded 422 vOTU clusters. Clusters detected in fewer than 5% of samples were subsequently removed, resulting in 417 vOTUs retained for downstream analysis. Each vOTU was classified into one of three lifestyle categories based on the annotation of its constituent member sequences as determined by CheckV^15^. vOTUs with all member sequences annotated as viruses, were classified as confirmed free viruses; all member sequences annotated as proviruses, were classified as confirmed proviruses and rest of the vOTUs containing a mixture of free virus and provirus member sequences were classified as mixed dual-lifestyle vOTUs. This classification yielded 359 confirmed free virus vOTUs, 48 confirmed provirus vOTUs, and 15 mixed dual-lifestyle vOTUs. Any gene (for AMR), vOTUs / PTUs (for viruses / plasmids), present in less than 3 samples was removed from the analysis.

Spearman rank correlation analysis was performed to assess co-occurrence patterns between bacterial species and antibiotic resistance gene (ARG) families. ARGs detected in ≥3 samples per group and bacterial species with a mean relative abundance ≥0.1% were included. Correlation coefficients (r) were calculated for all bacteria-ARG pairs, and significant positive associations (raw *p* < 0.01) were visualized as clustered heatmaps using the pheatmap package in R. Given the exploratory nature of the analysis and limited sample size (*n* = 7–8 per group), raw *p*-values were used, although no associations remained significant after BH-FDR correction.

### Statistical analysis

For abundance analysis, taxonomic composition and pathway analyses were performed using the microeco R package, with relative abundance values used as input. For AMR gene family and MGE analyses, raw read counts were normalized to Reads Per Kilobase per Million mapped reads (RPKM), calculated as read counts per contig normalized by contig length and the total number of sequenced reads per sample to account for differences in feature length and sequencing depth. Differences in relative abundance among groups were assessed using the Kruskal-Wallis rank-sum test for overall comparisons, followed by Dunn’s post hoc test for pairwise comparisons using the FSA R package. Resulting p-values were adjusted for multiple testing using the Benjamini–Hochberg false discovery rate (BH-FDR) method.

For microbial taxonomic composition and gut microbial pathway analyses, alpha-diversity metrics were calculated using the microeco R package. For AMR gene family and MGE analyses, diversity indices were calculated from relative abundance matrices using the vegan R package. Differences in alpha diversity among groups were assessed using the Kruskal-Wallis rank-sum test, followed by Dunn’s post hoc test, with p-values adjusted using the BH-FDR method.

For beta-diversity analysis, Bray-Curtis dissimilarity matrices were calculated from relative abundance data and visualized using principal coordinates analysis (PCoA). For microbial taxonomic composition and gut microbial pathway analyses, PCoA was performed using the microeco R package. For AMR gene family and MGE analyses, Bray-Curtis dissimilarity matrices were calculated using the vegdist() function and PCoA was performed using the cmdscale() function in the vegan R package. Overall differences in community composition among groups were assessed using permutational multivariate analysis of variance (PERMANOVA) implemented through the adonis2() function in the vegan package with 9,999 permutations. Because longitudinal samples were collected from the same individuals, permutations were stratified by participant (strata = individual) to account for repeated measures. Pairwise PERMANOVA analyses were performed using a paired design with participant stratification for within-cohort longitudinal comparisons (e.g., P_M0 vs P_M1, P_M0 vs P_M6, P_M0 vs P_M8, and H_M0 vs H_M6, etc). In contrast, comparisons between patient and healthy cohorts (e.g., P_M0 vs H_M0, etc) were conducted using an unpaired design without stratification, as these involved different individuals. For paired comparisons, analyses were restricted to participants with samples available at both time-points under comparison. Pairwise PERMANOVA p-values were adjusted using the BH-FDR method. Homogeneity of multivariate dispersions was additionally assessed using the betadisper() function with 9,999 permutations to complement the PERMANOVA results.

Exploratory differential features were identified using Linear Discriminant Analysis Effect Size (LEfSe) implemented in the microeco R package, with a significance threshold of raw p < 0.05. For species-level microbial signatures, an LDA score threshold ≥ 3.0 was applied, whereas an LDA score threshold ≥ 2.5 was used for functional pathway features derived from HUMAnN output. For bacterial-metabolites interactome analysis, the LDA identified exploratory taxa were retained to enable a more comprehensive evaluation of biologically meaningful microbial-metabolite interactions without compromising analytical robustness.

### Untargeted Fecal Metabolomics

Stool samples were lyophilized and aseptically pulverized. A 30 mg aliquot of each sample was homogenized in 500 µL of chilled methanol by vortexing for 10 min, followed by water-bath sonication for 30 min. After an additional 5 min vortex, the mixture was centrifuged (10,000 rpm, 15 min, 4°C). The supernatant was transferred to a clean tube, vacuum-dried for 3–4 h, and stored at −80°C. Reconstitution was performed in 300 µL of 50% methanol, followed by a 5 min vortex and 10 min sonication. After a final 1 min vortex and centrifugation (10,000 rpm, 10 min, room temperature), 200 µL of the supernatant was transferred to LC-MS vials. A pooled Quality Control (QC) sample was prepared by combining 10 µL from each sample vial, and 50% methanol was employed as the blank. Chromatographic separation was performed on a Thermo UHPLC Ultimate 3000 system utilizing two complementary modes at a column temperature of 40°C and an injection volume of 5 µL-1) Reverse-Phase Chromatography (RPC): Non-polar metabolites were separated using an ACQUITY HSS T3 column (2.1 mm × 100mm × 1.8μm, Waters). Mobile phase A consisted of 0.1% formic acid (FA) in water, and mobile phase B was 0.1% FA in methanol. The 14 min gradient at a flow rate of 300 µL/min was optimized as follows: 1% to 15% B (1 min), 15% to 35% B (3 min), 35% to 95% B (3 min), 95% B maintenance (2 min), and equilibration back to 1% B (1 & 4 min). 2) Hydrophilic Interaction Liquid Chromatography (HILIC): Polar metabolites were resolved using an XBridge BEH Amide column (2.1 mm x100mm x2.5μm, Waters). Mobile phase A comprised 20 mM ammonium acetate in water:acetonitrile (19:1 v/v, pH 9.0) with ammonium hydroxide, and mobile phase B was 100% acetonitrile. The 23 min elution gradient at a flow rate of 200 µL/min proceeded as follows: 85% to 30% B (3 min), 30% to 2% B (9 min), 2% B maintenance (3 min), and equilibration back to 85% B (1 & 7 _min)16,17._

Mass spectrometry was conducted on a high-resolution Q-Exactive platform in data-dependent MS2 (ddMS2) mode across positive and negative electrospray ionization (ESI) modes. MS1 scans were acquired over an m/z range of 65–1,000 da at a resolution of 120,000, and MS2 scans at 30,000. Source parameters included a vaporizer temperature of 310°C, capillary temperature of 300°C, sheath gas flow of 42 arb, auxiliary gas flow of 11 arb, and spray voltages of 3.5 kV (ESI+) and 3.0 kV (ESI−). Signal drift and mass accuracy were monitored by injecting a pooled QC sample after every five experimental runs.

### Data Preprocessing, Feature Annotation and Statistical and Bioinformatic Analysis

Raw LC-MS data processing was executed using the tidymass R package (v2.0.10). Peak detection was performed with a mass accuracy threshold of 30 ppm and a peak width range of 5–30 s.

Following feature alignment, QC-based signal correction and data cleaning were applied to filter out low-quality and noisy features. Missing values were imputed via the K-Nearest Neighbors (KNN) algorithm prior to data normalization and integration. Metabolite annotation was achieved against in-house and public MS/MS reference libraries using strict tolerances of 15 ppm for m/z and 30 s for retention time.

Downstream analysis was conducted using MetaboAnalyst 6.0^18,19^. Following data integrity verification, peak intensity data were normalized by total sum, log10 transformed, and Pareto-scaled. Interactive Principal Component Analysis (PCA) was applied to model and visualize overall metabolic variation across the study cohorts. Group-specific significance testing was performed using linear models without covariate adjustments, utilizing the P_M0 group as the baseline reference.

Differential exploratory metabolite features utility was evaluated using classical univariate Receiver Operating Characteristic (ROC) curves across designated binary and multi-group contrasts (H_M0 vs H_M6; H_M0 vs P_M0; P_M0 vs P_M6; P_M8 vs H_M6; and H_M0 vs P_M0 vs P_M6). Hierarchical cluster heatmaps of either the fully processed total data matrix or the statistically significant features were constructed using Heatmapper2 under default parameters.

Annotated metabolites were uploaded to the MetaboAnalyst Pathway Analysis module to retrieve their corresponding database identifiers. The canonical SMILES for each metabolite were then obtained and verified using PubChem. For metabolites without a directly available SMILES entry, the compound name was searched in PubChem to retrieve the corresponding canonical SMILES. The validated canonical SMILES were subsequently used for chemical classification at the superclass level using the online ClassyFire platform.

### Multi-Omics Data Integration and Interactomics

The microbiome and metabolome features identified by the LEfSe (for 5 different group comparisons) and univariate Linear modeling, respectively, were selected and grouped to make two unique lists of 61 bacterial taxa and 50 metabolite features. Both input data were log10-transformed. These differential metabolites and microbial taxa at species levels were correlated using the mixOmics package (v6.32.0)^20^ on R (version 4.5.0). Multi-omics integration was conducted using the DIABLO (Data Integration Analysis for Biomarker discovery using Latent Components) framework, implemented in the mixOmics package via the block.splsda() function. To evaluate pairwise associations between 50 metabolites and 61 microbes^21^, Spearman correlation tests were conducted for all possible metabolite-microbe pairs using cor.test() with method = “spearman”. For each pair, the Spearman rho coefficient and its associated p-value were recorded. Benjamini-Hochberg False Discovery Rate (BH-FDR) correction^22^ was then applied across all pair-wise p-values to control for the multiple comparison burden arising from the combinatorial expansion of feature pairs. A global microbe-metabolite Spearman correlation matrix of 3050 initial potential correlations, representing all targeted metabolites and microbes was generated. High-confidence interactomic links were identified using a stringent three-tier filter requiring the metabolite, the microbial taxon, and their pairwise Spearman correlation to each independently achieve a BH-FDR < 0.05. Only pairs satisfying all three criteria simultaneously were designated as final significant pairs which included 19 unique microbial species correlated with 16 unique functional metabolites, to generate 56 true cross-omics significance edges. Heatmap was generated using pheatmap (version 1.0.13)^23^ to visualize the network of statistically significant microbiome-metabolome associations.

## RESULTS

### 1) Basic characteristics of the study cohort

Eight patients with culture-confirmed pulmonary tuberculosis, along with their healthy household close contacts (n=8) aged 18-60 years residing in southern India, were enrolled in the study (Fig. 1A). Participants with other lung disease, systemic or immune-compromising disease, smoking, alcohol use, along with recent medication exposure (only for the control group) were excluded from the study. Age did not differ significantly between the cohorts (unpaired t-test, t (14) = 0.095, p = 0.925), whereas baseline BMI was lower in patients than controls (p = 0.0015; Table 1). Longitudinal evaluation showed no significant changes in BMI within the healthy cohort over time (H_M0 vs. H_M6; paired t-test, p = 0.4617). However, the overall longitudinal effect on BMI within the patient cohort was marginally significant (mixed-effects REML, p = 0.0780), along with notable upward trends observed between baseline and subsequent post-treatment intervals i.e. from baseline to month 6 (p = 0.0007) and month 8 (p = 0.0011, Table 1). The Complete Blood Count (CBC) parameters remained stable in controls from H_M0 to H_M6 (p > 0.05). At baseline, controls and patients differed only in platelets (p = 0.014), ALC (p = 0.024), and AMC (p = 0.013). In patients, mixed-effects REML analysis showed significant treatment effects for hemoglobin, PCV, WBC, platelets, ANC, and AMC (p < 0.05), driven mainly by later time-point changes, highlighting significant declines in WBC and ANC at months 6 and 8 relative to baseline. RBC, MCV, MCH, MCHC, AEC, ALC, and RDW did not change significantly. Absolute basophil count (ABC) analysis was excluded due to frequent missing values (Table 1).

**Figure 1.**
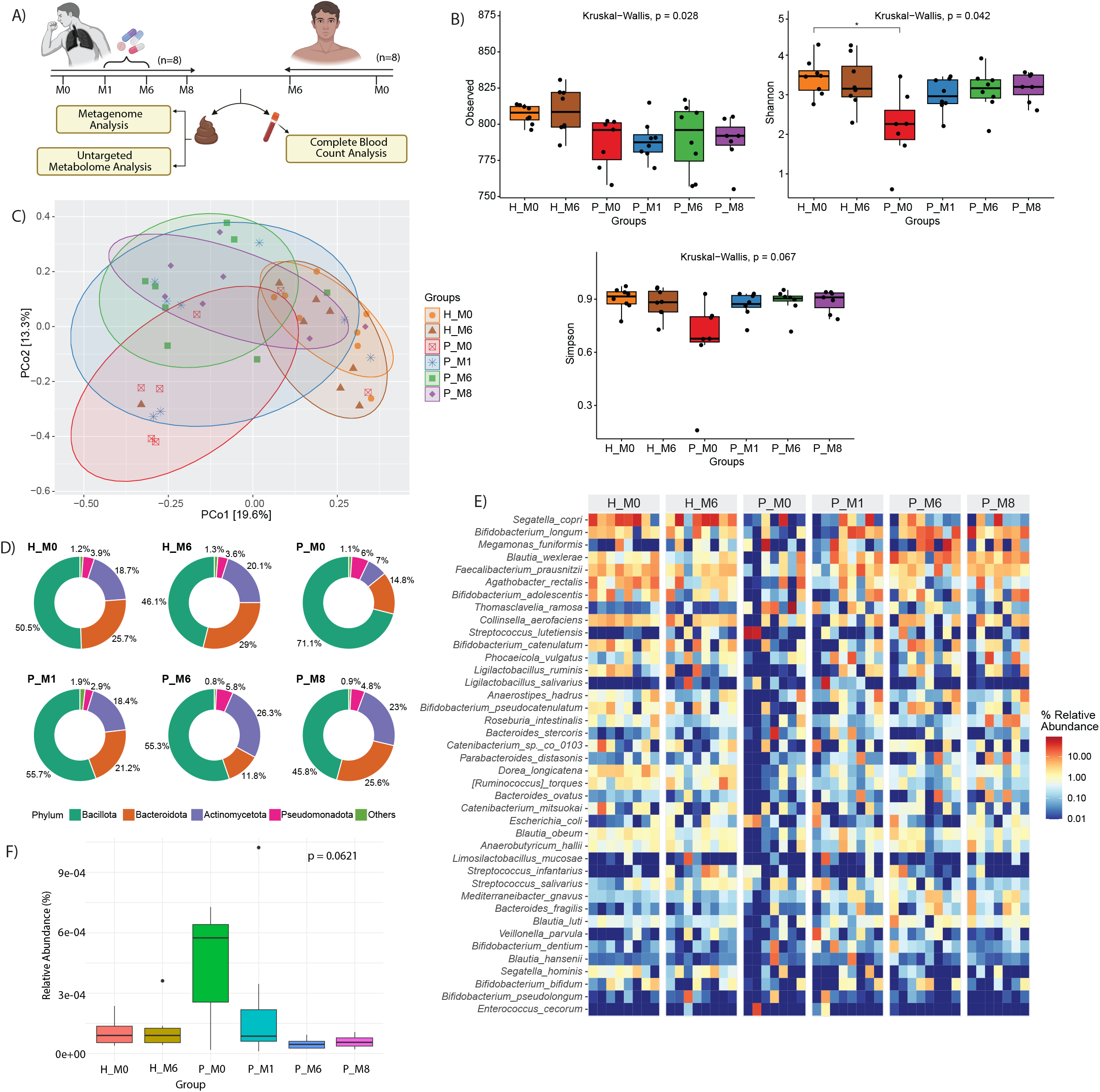
Gut microbial diversity and taxonomic composition are altered in tuberculosis patients and progressively recover during anti-tuberculosis treatment. (A) Study design (*created using BioRender*) (B) Alpha diversity metrics (Observed, Shannon, and Simpson indices) comparing healthy controls (H_M0, H_M6) and tuberculosis patients (P_M0, P_M1, P_M6, and P_M8) across longitudinal sampling time-points. (C) Bray-Curtis PCoA showing separation of baseline TB microbiota from healthy controls, with partial convergence during treatment. (D) Average phylum-level microbial composition demonstrating temporal shifts in the relative abundance of Bacillota, Bacteroidota, Actinomycetota, Pseudomonadota, and other taxa. (E) Heatmap depicting the relative abundance of the most abundant bacterial species across all samples. (F) Relative abundance of *Mycobacterium tuberculosis* in fecal metagenomes across all time-points. Boxplots show median and interquartile range with individual samples overlaid. Analyses were done using the Kruskal–Wallis test. Exact global p-values are shown; asterisks indicate pairwise significance.

**Table 1:**
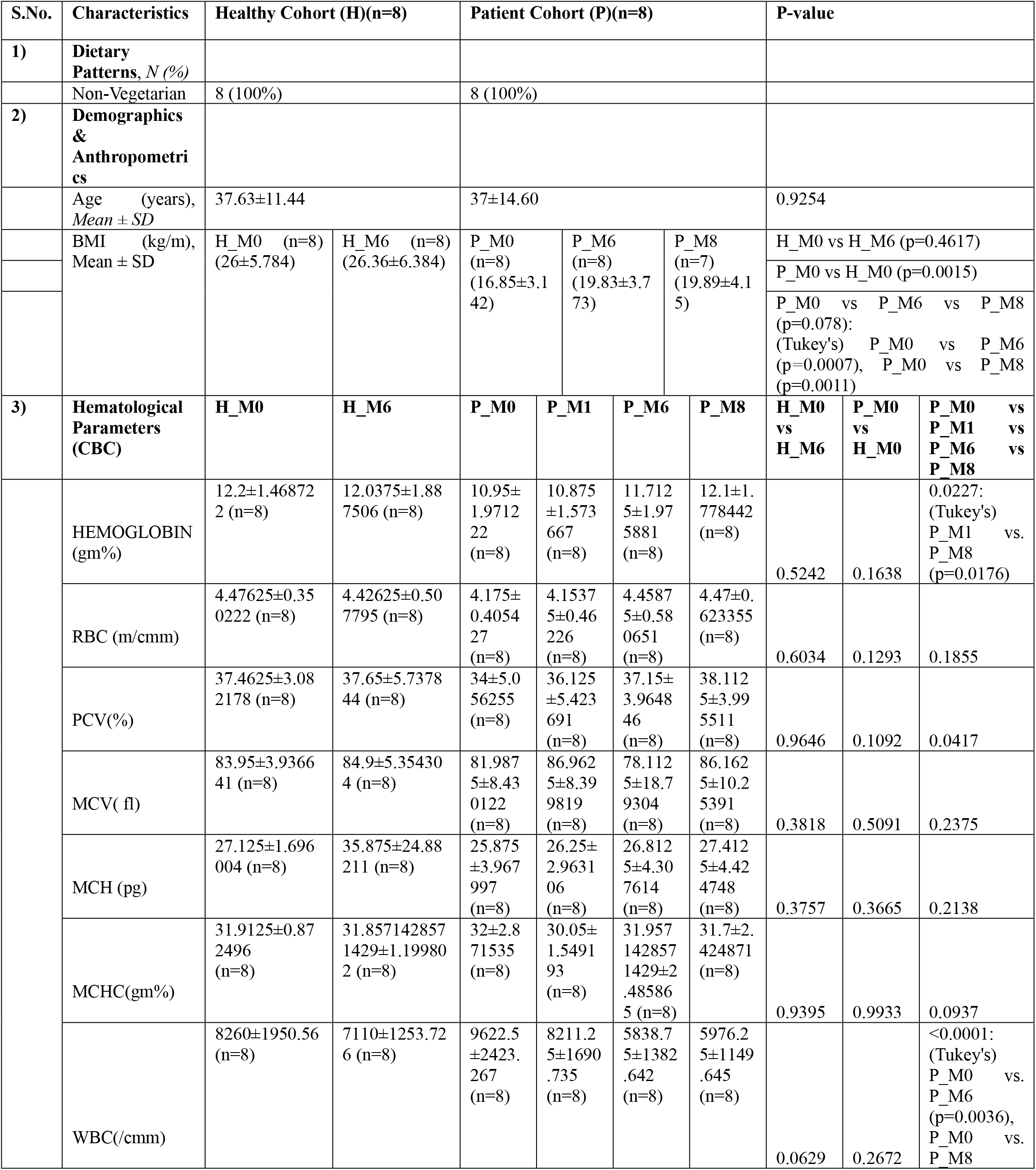
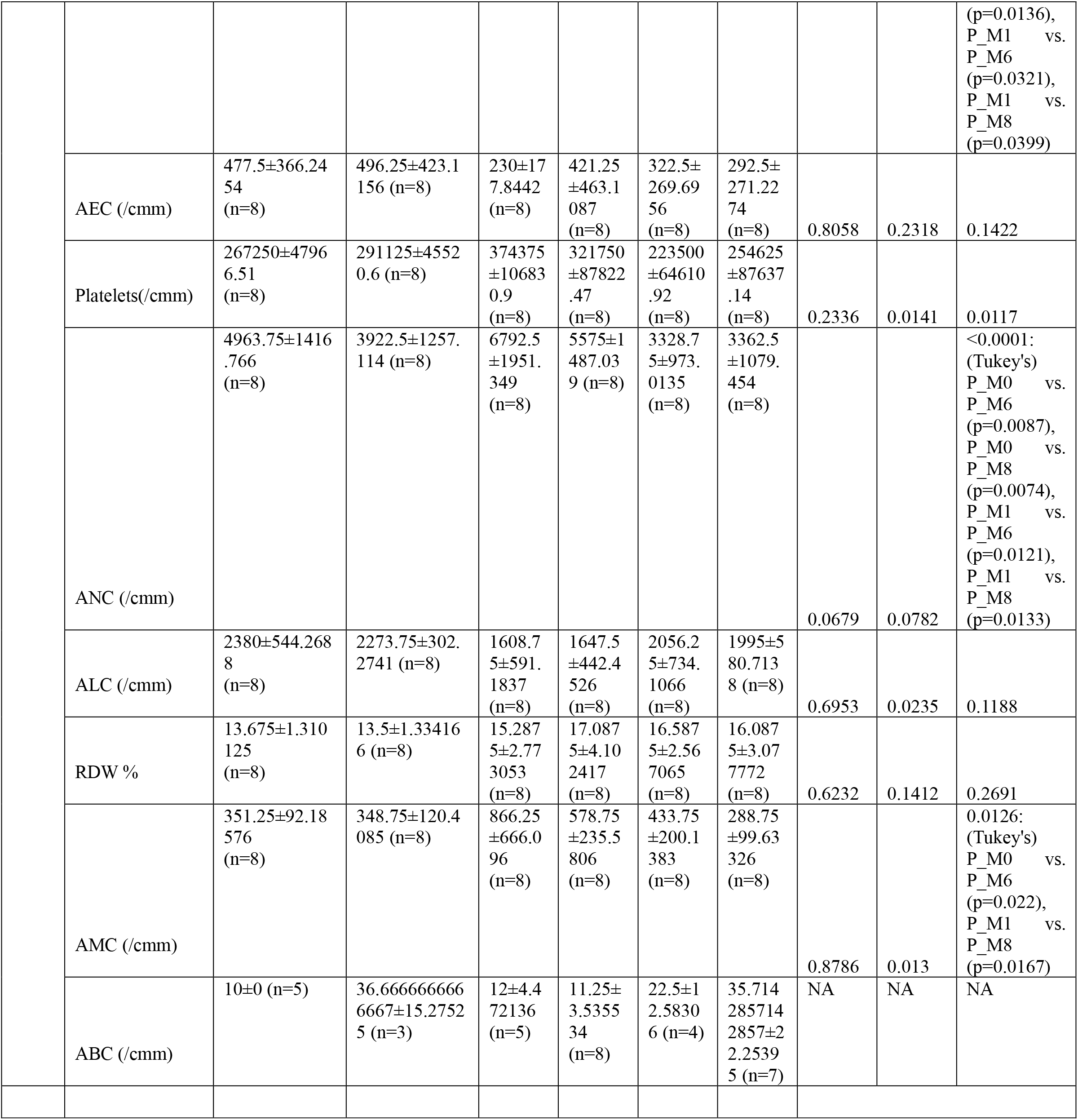
Baseline Demographic and Clinical Characteristics of Participants.

### 2) Pre-treatment Characterization of the Intestinal Microbiota in Tuberculosis Patients

We assessed whether pulmonary *Mycobacterium tuberculosis* (Mtb) infection was associated with altered intestinal microbiota by comparing baseline shotgun metagenomic sequencing profiles of treatment-naive pulmonary tuberculosis patients (P_M0, n = 7) with the healthy household contacts (H_M0, n = 8).

Treatment-naive patients (P_M0) showed significantly lower Shannon diversity (p_adj = 0.033) and Simpson diversity (p_adj = 0.06) compared to H_M0, while observed OTUs (p_unadj = 0.025; p_adj = 0.12) remained nominally lower but non-significant after multiple test correction (Fig. 1B). The variations in global gut microbial community structure were analyzed by Bray-Curtis distances using PERMANOVA which showed an overall difference in microbial composition across clinical groups (R2 = 0.171, F = 1.65, p = 0.049), with a significant baseline shift between P_M0 and H_M0 after BH correction (R2 = 0.185, p = 0.0053, padj = 0.039). Since global Beta-dispersion differed across groups (F = 4.5, p = 0.002), we assessed pairwise beta-dispersion and found a significant difference at baseline (F = 25.63, p = 0.0001, padj = 0.0015; Fig. 1C).

We next compared microbiome composition to identify taxa differing between groups. In P_M0, the Bacillota:Bacteroidota ratio was markedly higher than in H_M0 (4.8 vs. 1.96), with reduced Actinomycetota (7% vs 18.7%) and a slight increase in Pseudomonadota (6% vs 3.9%) (Fig. 1D). At the species level, P_M0 showed reduced abundance of common gut commensals, including *Segatella copri*, several *Bifidobacterium* species, *Faecalibacterium prausnitzii*, *Collinsella aerofaciens*, *Ligilactobacillus ruminis*, *Catenibacterium* species*, Parabacteroides distasonis*, *Dorea longicatena*, *Ruminococcus torques*, *Bacteroides ovatus*, *Blautia* species, and *Limosilactobacillus mucosae*, involved in complex carbohydrate fermentation and SCFA production, with several also contributing to immune regulation, intestinal barrier maintenance, and host metabolic homeostasis^24,25^. In contrast, P_M0 was enriched for many opportunistic pathobionts associated with disrupted or inflamed guts, including *Thomasclavelia ramose*, *Streptococcus* species, *Bacteroides stercoris*, *B. fragilis*, *Escherichia coli*, *Megamonas funiformis*, *Bifidobacterium dentium*, *B. pseudolongum*, *Ligilactobacillus salivarius*, and *Blautia hansenii* (Fig. 1E).

### 3) Effect of ATT on Gut Microbiota Composition Over Time

We next assessed the longitudinal effects of ATT on the gut microbiota of TB patients. Alpha diversity increased gradually after treatment initiation, although differences from P_M0 were not significant after adjustment (Dunn’s test, padj > 0.05). Nominal increases were observed for Shannon (P_M6: p_unadj = 0.052 & P_M8: p_unadj = 0.017), and Simpson (P_M6: p_unadj = 0.017 & P_M8: p_unadj = 0.012) indices (Fig. 1B).

Global Bray-Curtis PERMANOVA showed significant overall differences in beta diversity across clinical groups (R^2^ = 0.171, F = 1.65, p = 0.049; Fig. 1C), but within patients, no significant longitudinal centroid shifts from P_M0 was observed after BH correction (padj ≥ 0.058), although P_M0 showed marginal differences from P_M6 (p_unadj = 0.015) and P_M8 (p_unadj = 0.031). Beta-dispersion revealed significant heterogeneity across groups (F = 4.56, p = 0.002), with P_M0 having significant pairwise dispersion with P_M8 (padj = 0.013) and marginal difference with P_M6 (p_unadj = 0.026); H_M0 also differed significantly from P_M1 (padj = 0.024). Together, these findings suggest that pulmonary Mtb infection is marked by baseline taxonomic disruption and increased inter-individual microbial instability, which gradually stabilizes by the end of the observed monitoring period.

Longitudinal taxonomic profiling showed dynamic recovery of the gut microbiota during and after ATT. At the phylum level, the Bacillota:Bacteroidota ratio decreased from P_M0 to P_M1 (4.8 vs 2.62), rose again at P_M6 (4.68), and approached control-like levels by P_M8 (1.78); Actinomycetota also increased over time, whereas Pseudomonadota was broadly comparable across groups except for a transient reduction at P_M1 (Fig. 1D). At the species level, several commensal and SCFA-associated taxa including *Bifidobacterium* species*, F. prausnitzii, Blautia* species, *Catenibacterium sp. co_0103, V. parvula, P. distasonis, L. ruminis, Bacteroides* species*, M. gnavus, A. hallii, C. aerofaciens, P. vulgatus* increased during treatment, while multiple opportunistic or dysbiosis-associated taxa which included *E. coli, Thomasclavelia ramose, S. lutetiensis, B. stercoris,* along with a few beneficial commensals like *B. dentium, B. hansenii, A. rectalis, R. intestinalis* declined relative to P_M0. By P_M8, most taxonomic profiles remained broadly comparable to P_M6, with only a subset showing further increases or decreases (Fig. 1E).

Interestingly, Mtb DNA was detectable in fecal WGS data, with the highest, though non-significant, abundance in treatment-naive patients (p = 0.0621). After ATT initiation, intestinal Mtb levels declined progressively (padj > 0.05), reaching minimal levels by months 6 (p_unadj = 0.004) and 8 (p_unadj = 0.018), indicating effective reduction of gut Mtb burden with therapy (Fig. 1F).

Taken together, these data show that treatment naive patients show maximum decline in microbiota diversity, while, even though under antibiotic treatment, the patients regain the bacterial diversity during treatment.

### 4) Longitudinal Assessment of the Compositional and Functional Alterations in the Gut Microbiota within Healthy Cohort

Longitudinal changes were assessed to determine the influence of household exposure to active Mtb patient on the gut microbiota of healthy close contacts. Pairwise alpha diversity, beta-diversity centroids (padj = 0.5022), and dispersion (padj = 0.310), along with phylum-level abundance indicated stability over six months in household contacts (Fig. 1B-D). However, subtle taxonomic fluctuations were visible across timepoints. At baseline, the comparative abundance of some obligate anaerobes and butyrate-producing genera, including *Blautia*, *Agathobacter*, *Bacteroides*, *Roseburia*, *Catenibacterium*, *Dorea*, and *Bifidobacterium* was slightly higher. By month 6, the abundance shifted towards specific facultative anaerobes and common gut residents, including *S. lutetiensis*, *L. salivarius*, *E. coli*, *L. mucosae*, *V. parvula*, *B. pseudolongum*, and *S. infantarius* (Fig. 1E). While compositional alterations were not significant, these taxonomic fluctuations indicated subtle, sub-clinical taxonomic restructuring occuring within household contacts longitudinally.

To determine the specific microbial identities and metabolic pathways driving structural variations across cohorts, we performed Linear Discriminant Analysis Effect Size (LEfSe) modeling (Tables S2 & S3). Despite maintaining stable community architectures, high-resolution LEfSe modelling revealed a significant temporal shift in specific taxonomic and functional features within healthy controls over six months. The H_M0 group was enriched with obligate anaerobic, SCFA-producing taxa, including *Dorea* species (*D. longicatena*, *D. formicigenerans*), *Roseburia intestinalis*, and *Faecalibacterium* spp. Functionally, this group was characterized by biosynthetic and nutrient recycling pathways, including chorismate biosynthesis I and the superpathway of adenosylcobalamin (vitamin B12) salvage, reflecting a metabolically versatile, homeostatic, and cooperative community. Conversely, at month 6, a profound expansion of facultative anaerobes and lactic acid bacteria, predominantly from the genus *Streptococcus* (*S. infantarius*, *S. lutetiensis*, *S. ruminicola*, *S. equinus*), alongside *Ligilactobacillus salivarius*, and *Veillonella atypica* was observed. Mirroring this taxonomic shift, the functional profile switched toward fermentative pathways linked to stress and redox balancing, specifically the superpathways of (R,R)-butanediol and 2,3-butanediol biosynthesis. This parallel restructuring uncovers a sub-clinical transition from a biosynthetically integrated ecosystem toward a fermentation-driven, dysbiotic state within the shared household environment over time (Fig. 2A & Fig. S1A).

**Figure 2.**
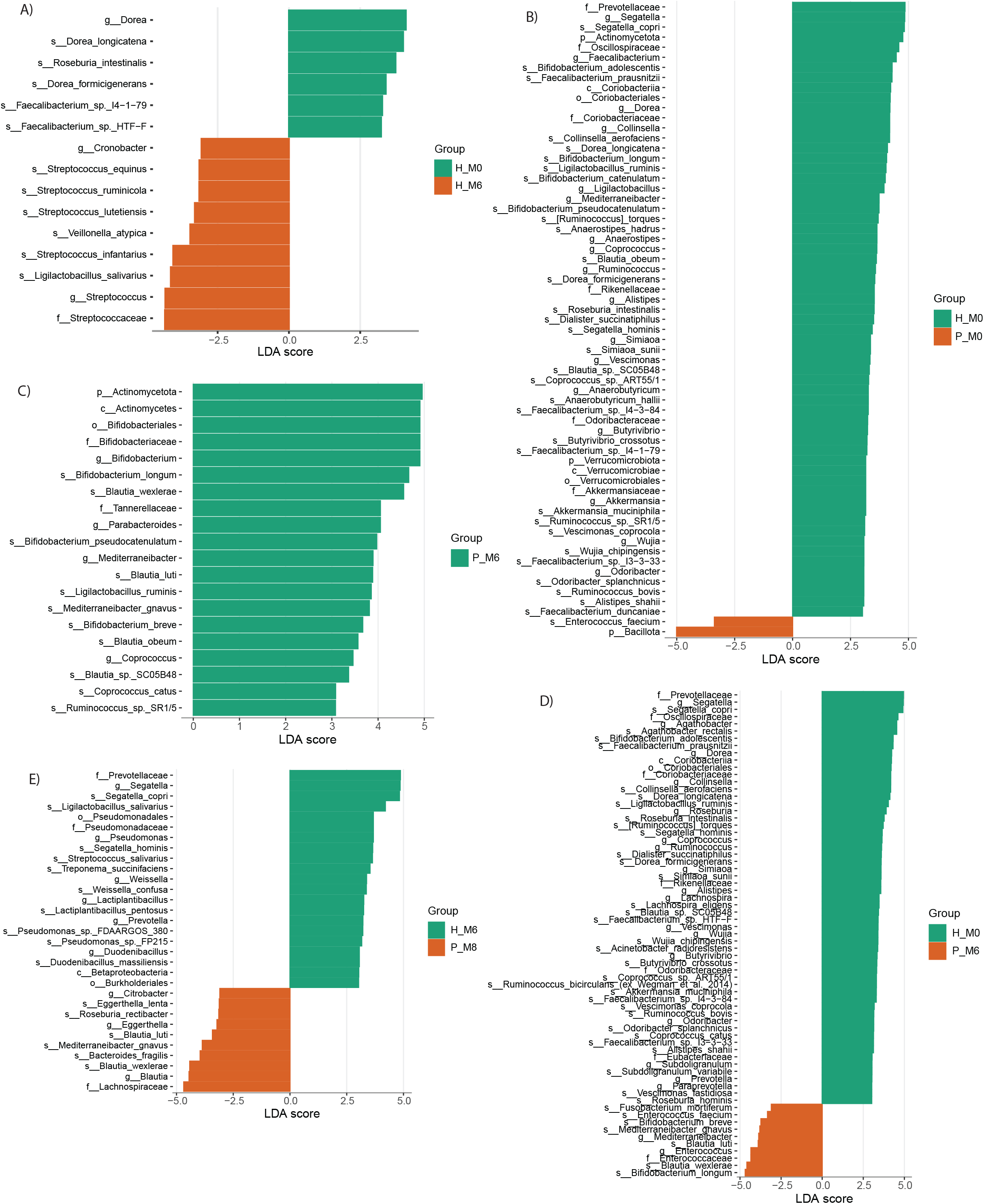
Identification of exploratory structural features associated with tuberculosis and anti-tuberculosis treatment by Linear Discriminant Analysis Effect Size (LEfSe) analysis. Linear Discriminant Analysis (LDA) effect size plots highlighting enriched microbial taxa across longitudinal baseline and follow-up comparisons between healthy controls (H) and patients (P). Plots display taxonomic features at species levels. Exploratory microbial taxa (LDA score > 3, p < 0.05) are highlighted according to group assignment. (A) Longitudinal baseline vs. follow-up in healthy controls (H_M0 vs. H_M6). (B) Baseline profiles depicting dysbiotic shifts between healthy controls (H_M0) and treatment-naive patients (P_M0). (C) Treatment-driven taxonomic shifts (P_M0 vs. P_M6). (D) Multigroup comparison (H_M0 vs. P_M0 vs. P_M6) mapping exploratory profiles across baseline healthy controls, baseline patients, and post-treatment patients. (E) Endpoint Comparison (H_M6 vs. P_M8): Plot identifying persistent variations between healthy follow-up controls (H_M6) and post-treatment follow-up patients (P_M8).

### 5) Longitudinal Assessment of the Compositional and Functional Alterations in the Gut Microbiota within Patient Cohort

The cross-sectional study at baseline using LEfSe modelling exposed a marked structural and functional asymmetry between healthy controls (H_M0) and treatment-naive patients (P_M0) (Fig. 2B & Fig. S1B). The H_M0 cohort demonstrated a pronounced enrichment of health-associated obligate anaerobes and SCFA producers (*Faecalibacterium prausnitzii*, *Roseburia intestinalis*, *Anaerostipes hadrus*, *Dorea* spp., *Blautia* spp.), beneficial Actinobacteria (*Bifidobacterium longum*, *B. adolescentis*, *Collinsella aerofaciens*), and specialized carbohydrate/mucin fermenters (*Akkermansia muciniphila*, *Alistipes shahii*, *Odoribacter splanchnicus*, *Segatella (S. copri*, *S. hominis)*, *Ruminococcus* spp. This taxonomic diversity aligned with a robust enrichment of anabolic pathways, including multi-amino acid biosynthesis (methionine, isoleucine, arginine, glutamine, and branched-chain amino acids), central carbon metabolism (glycolysis, gluconeogenesis), and essential cofactor/vitamin pathways (folate transformations, thiamine diphosphate biosynthesis, NAD, and vitamin B12 salvage). In stark contrast, the P_M0 cohort exhibited a profound collapse of both taxonomic and functional diversity. Structurally, only *Enterococcus faecium*, an opportunistic pathobiont associated with clinical dysbiosis, was identified as a robust discriminator. Functionally, P_M0 was restricted to a narrow set of catabolic and stress-associated pathways, including purine ribonucleoside degradation, nitrate reduction, and butanediol biosynthesis. These findings indicate that acute, untreated pulmonary Mtb infection is coupled with a highly depleted, functionally constrained gut microbiome characterized by a loss of critical biosynthetic machinery.

Longitudinal tracking within the patient cohort revealed a stepwise restoration of both community composition and metabolic competence following the initiation of ATT. Upon comparing P_M0 to P_M6, while the highly volatile acute phase (P_M0) showed restricted functional enrichment (seleno-amino acid and L-arginine biosynthesis), the month 6 treatment group (P_M6) experienced a massive expansion across multiple metabolic domains. This functional reactivation was driven by a strong taxonomic resurgence of probiotic-associated *Bifidobacterium* species (*B. longum*, *B. breve*, *B. pseudocatenulatum*) and SCFA-producing Lachnospiraceae (*Blautia wexlerae*, *B. luti*, *B. obeum*, *Coprococcus catus*, *Subdoligranulum variabile*, *Ruminococcus* spp.). Functionally, the P_M6 ecosystem expanded its capacity for central energy metabolism (glycolysis, pyruvate fermentation to acetate/lactate), nucleotide biosynthesis (pyrimidine ribonucleotides/deoxyribonucleotides), amino acid synthesis (lysine, threonine, methionine), and carbohydrate/glycan utilization, supplemented by thiamine diphosphate salvage and the γ-glutamyl cycle (Fig. 2C & Fig. S1C).

Next, the stepwise multi-group trajectory comparing control (H_M0), active infection group (P_M0) and treatment completion group (P_M6) confirmed a distinct stratification of metabolic and taxonomic capacity. The H_M0 group occupied the upper tier of metabolic complexity, exhibiting extensive enrichment in biosynthetic, cell structure (peptidoglycan), and translational (tRNA charging) pathways. The P_M6 cohort occupied a transitional, intermediate state; it demonstrated clear signs of functional restoration, particularly in carbohydrate metabolism (starch biosynthesis, glycogen degradation), fatty acid biosynthesis, and glycan degradation, yet retained signatures of incomplete recovery through the persistent enrichment of opportunistic pathobionts (*Enterococcus faecium* and *Fusobacterium mortiferum*). Crucially, the P_M0 group sat at the bottom of the cascade, showing minimal enrichment outside of arginine biosynthesis and butanediol fermentation (Fig. 2D & Fig. S1D).

To understand the resolution and adaptation at month 8, we compared P_M8 to H_M6. The divergence between prolonged household exposure and therapeutic patient recovery was fully realized by month 8. Surprisingly, while the exposed H_M6 group remained locked in a dysbiotic, functionally constrained state characterized by opportunistic *Pseudomonas* spp. and *Weissella confusa*, *Streptococcus* species, peptidoglycan biosynthesis, and NAD salvage, the treated P_M8 cohort achieved advanced functional stabilization. The P_M8 microbiome was enriched with obligate anaerobic commensals (*Blautia wexlerae*, *B. luti*, *R. rectibacter*, *M. gnavus*, *B. fragilis*, *E. lenta*). This taxonomic stabilization was tightly coupled with enhanced metabolic flexibility and ecological adaptation, featuring a rich abundance of pathways for amino acid/cofactor biosynthesis (L-cysteine, thiamine, biotin), central carbon utilization (pentose phosphate and Entner–Doudoroff pathways), and advanced carbohydrate/glycan degradation (starch, GDP-mannose, N-acetylglucosamine, neuraminate) (Fig. 2E & Fig. S1E).

Taken together, these results demonstrate a clear, coordinated trajectory where the profound structural and functional dysbiosis induced by active Mtb infection is gradually overcome by long-term systemic ATT, guiding the gut ecosystem toward metabolic diversification and ecological stabilization by month 8.

### 6) Fecal Metabolomic Profiling Across Disease Progression and Therapeutic Phases

To determine if the taxonomic and functional alterations corresponded to biochemical changes within the intestinal lumen, untargeted fecal metabolomic profiling was performed across all study groups (Table S4). Following data filtering, 582 metabolites were retained for analysis. Hierarchical clustering analysis (HCA) revealed distinct group-specific metabolomic signatures that closely mirrored the metagenomic findings. The global dendrogram separated samples into two principal clusters, with healthy household contacts (H_M0 and H_M6) clustering together and all patient groups (P_M0, P_M1, P_M6, and P_M8) forming a separate branch (Fig. S2A).

Principal component analysis (PCA) further demonstrated significant differences in global metabolomic profiles among study groups (PC1 = 12.9%, PC2 = 8.4%; PERMANOVA: F= 3.953, R²= 0.33, p = 0.001, 999 permutations), with the study groups collectively accounting for approximately 33% of total metabolomic variation (Fig. 3A). Healthy controls (H_M0 and H_M6) clustered closely together, whereas treatment-naive patients (P_M0) occupied a distinct and more dispersed region of the PCA space, indicating greater inter-individual metabolic variability. Following ATT initiation, patient samples exhibited a gradual temporal shift, with P_M1 and P_M6 occupying intermediate positions that partially overlapped with P_M0. By month 8 (P_M8), samples formed a more compact cluster distinct from baseline disease but remained spatially separated from healthy controls, indicating partial, but incomplete, restoration of the gut metabolome.

**Figure 3.**
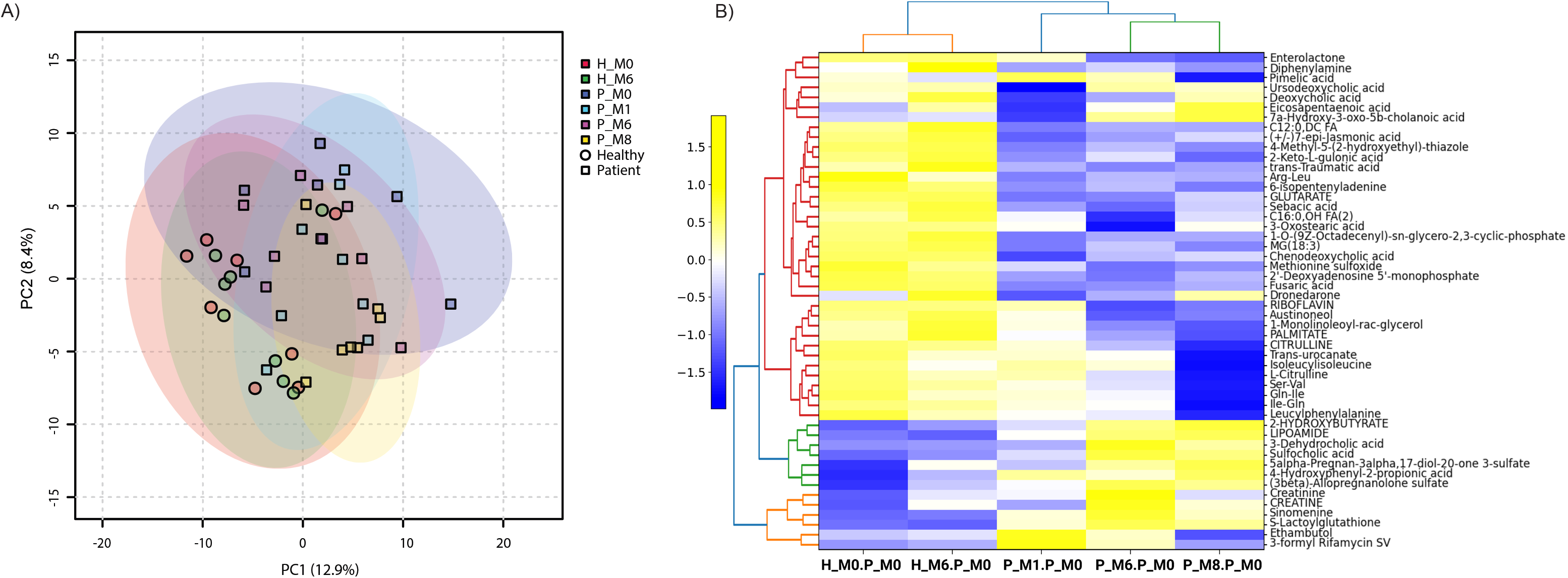
Untargeted Stool Metabolomics Profiling and Differential Exploratory Features Identification. (A) Principal Component Analysis (PCA): Scores plot depicting global metabolic variance between healthy controls and patients across time points (PC1 = 12.9%, PC2 = 8.4% variance explained). (B) Differentially Abundant Metabolites (Linear Modeling Analysis): Hierarchical clustering heatmap of statistically significant stool metabolites normalized to patient baseline (P_M0).

This longitudinal metabolomic trajectory closely recapitulated the microbial community dynamics observed by beta-diversity analysis (Fig. 1C), with progressive movement away from the acute disease state during treatment while remaining distinct from the healthy gut ecosystem at month 8. Together, these findings demonstrate that prolonged ATT is associated with substantial metabolic recovery but does not completely restore the intestinal metabolome to the healthy state within the study period.

To identify metabolites associated with disease progression and recovery, univariate linear models with false discovery rate correction (FDR < 0.05) were applied using treatment-naive patients (P_M0) as the reference group. This analysis identified 50 significantly altered metabolites, predominantly belonging to the lipid and lipid-like molecules and organic acids and derivatives superclasses (Fig. 3B & Fig. S2B; Table S5). Hierarchical clustering of these metabolites further highlighted the metabolic distinction between disease, healthy or recovering states.

Healthy controls were characterized by enrichment of homeostatic host-microbiome co-metabolites, including the secondary bile acids ursodeoxycholic acid, deoxycholic acid, and 7α-hydroxy-3-oxo-5β-cholanoic acid, together with amino acids, dipeptides, and vitamins such as Ser-Val, Gln-Ile, L-citrulline, and riboflavin, which were reduced during the early stages of treatment. In contrast, early treatment patient group (P_M1) were enriched with stress-associated metabolites and anti-tuberculosis drug markers, including ethambutol and 3-formyl rifamycin SV. During and following treatment (P_M6 and P_M8), the metabolome showed progressive enrichment of metabolites associated with cellular energy metabolism and organic acid metabolism, including 2-hydroxybutyrate, 3-dehydrocholic acid, sulfocholic acid, and creatinine (Fig. 3B). These metabolomic changes provide a chemical validation of the functional microbiome recovery observed in metagenomic data.

### 7) Exploratory Metabolite Signatures and Multi-Omics Interaction Network

To explore metabolite features capable of distinguishing between study groups, classical receiver operating characteristic (ROC) curve analysis was performed using the MetaboAnalyst Biomarker Analysis module for five comparisons (H_M0 vs H_M6, P_M0 vs H_M0, P_M0 vs P_M6, P_M8 vs H_M6, and H_M0 vs P_M0 vs P_M6). Applying filtering criteria of area under the curve (AUC ≥ 0.70) and nominal statistical significance (p ≤ 0.05), we identified candidate discriminatory metabolites for each comparison. (Table S6). Compiling these features from all five comparisons, yielded a panel of 222 metabolites. Notably, all 50 FDR corrected significantly differential metabolites identified previously were retained within this ROC-validated framework.

To investigate interactions between microbial and metabolic alterations, the 50 significantly differential metabolites were integrated with the 61 LEfSe-derived exploratory taxonomic signatures identified across the same comparisons. Multi-omics integration was performed using the Multilevel_DIABLO framework in mixOmics, followed by FDR filtering of microbial features (Microbe_FDR ≤ 0.05), metabolite features (Metabolite_FDR ≤ 0.05), and Spearman correlations (Spearman_FDR_0.05 = True) (Table S7). From 3050 microbe-metabolite associations, this workflow identified 56 significant interactions (Final_significant = True), linking 19 microbial species with 16 metabolites (Table S8).

Multi-omics integration revealed coordinated associations between candidate discriminatory microbial taxa and metabolites (Fig. 4). Bile acid metabolism, protein catabolism and ATT drug and derivatives constituted prominent interaction axes. The metabolites like 2-hydroxybutyrate, bile acid metabolites, lipoamide, and (3β)-allopregnanolone sulfate showed positive associations with several beneficial anaerobic gut commensals, while negative associations with *Pseudomonas* spp. (FP215 and FDAARGOS_380). In contrast, the dipeptides and amino acids have exhibited positive associations with these *Pseudomonas* spp. And negative associations with gut commensals. The ATT drug Ethambutol showed negative correlation with *B. breve and E. lenta* while positive association with *L. salivarius.* The rifampicin derivative 3-formyl Rifamycin SV had a negative correlation with *E. lenta.* Collectively, these interaction patterns demonstrate coordinated relationships between microbial dysbiosis, altered protein absorption and bile acid metabolism, oxidative stress, and metabolites associated with intestinal barrier function during active TB and recovery following ATT.

**Figure 4.**
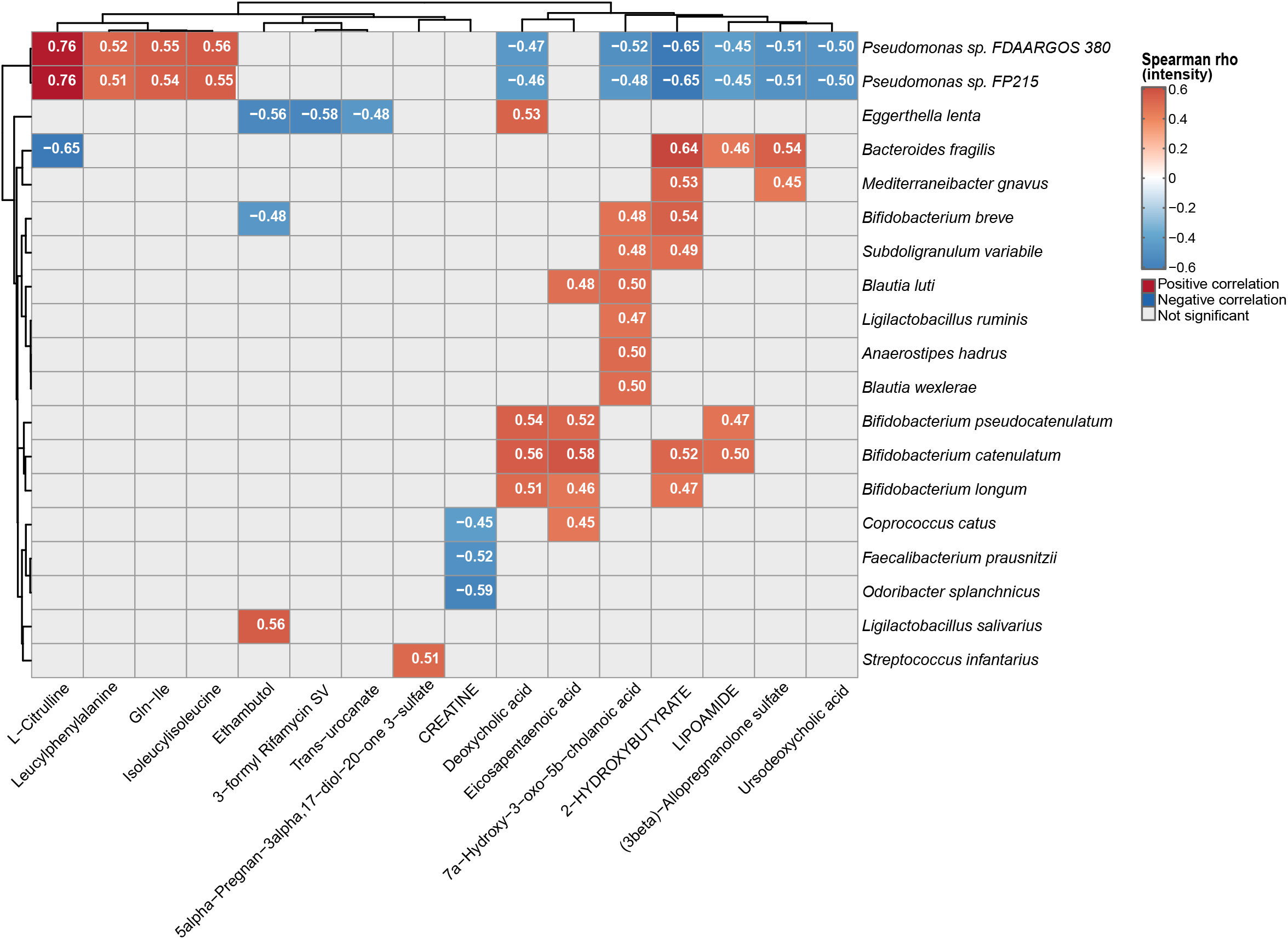
Multi-Omics Interactome Mapping Gut Microbiota-Metabolite Cross-Talk. Heatmap showing the integrated network constructed via Multilevel_DIABLO (mixOmics) linking LEfSe identified exploratory microbial signatures and LM identified fecal metabolites. The core interactome displays 56 significant edges connecting 19 microbial species with 16 functional metabolites, filtered by dual-feature and Spearman correlation FDR constraints (FDR ≤ 0.05). Color gradient indicate positive (red) and negative (blue) associations between taxa and metabolites.

### 8) Longitudinal Dynamics and Resilience of the Gut Resistome

Functional impact of Mtb pathogenesis and subsequent ATT on the intestinal resistome was analyzed by longitudinal gut resistome alpha diversity (Fig. 5A). Although global diversity did not significantly differ across groups (Kruskal-Wallis; Obs.: p = 0.277, Shan.: p = 0.135, Simp.: p = 0.082), exploratory pairwise comparisons revealed subtle, suggestive longitudinal trends. Healthy baseline controls (H_M0) maintained the lowest overall antimicrobial resistance gene (ARG) richness and diversity, with a modest increase by month 6 (H_M6). Conversely, patients consistently displayed elevated resistome diversity metrics. This divergence was initially evident at the untreated baseline (P_M0 vs. H_M0: Shan. p_unadj = 0.022, Simp. p_unadj = 0.026), pointing to a disease-associated expansion of resistance-carrying lineages. Intermediate ATT phases (P_M1 and P_M6) exerted only a transient, moderate suppression of diversity, which rebounded by post-treatment monitoring (P_M8 vs. H_M0: Obs. p_unadj = 0.043, Shan. p_unadj = 0.018, Simp. p_unadj = 0.004). While these pairwise variations did not survive FDR correction (p_adj > 0.05), they pointed to a localized, longitudinal expansion of the patient AMR reservoir relative to unexposed baseline controls (Fig. 5A).

**Figure 5.**
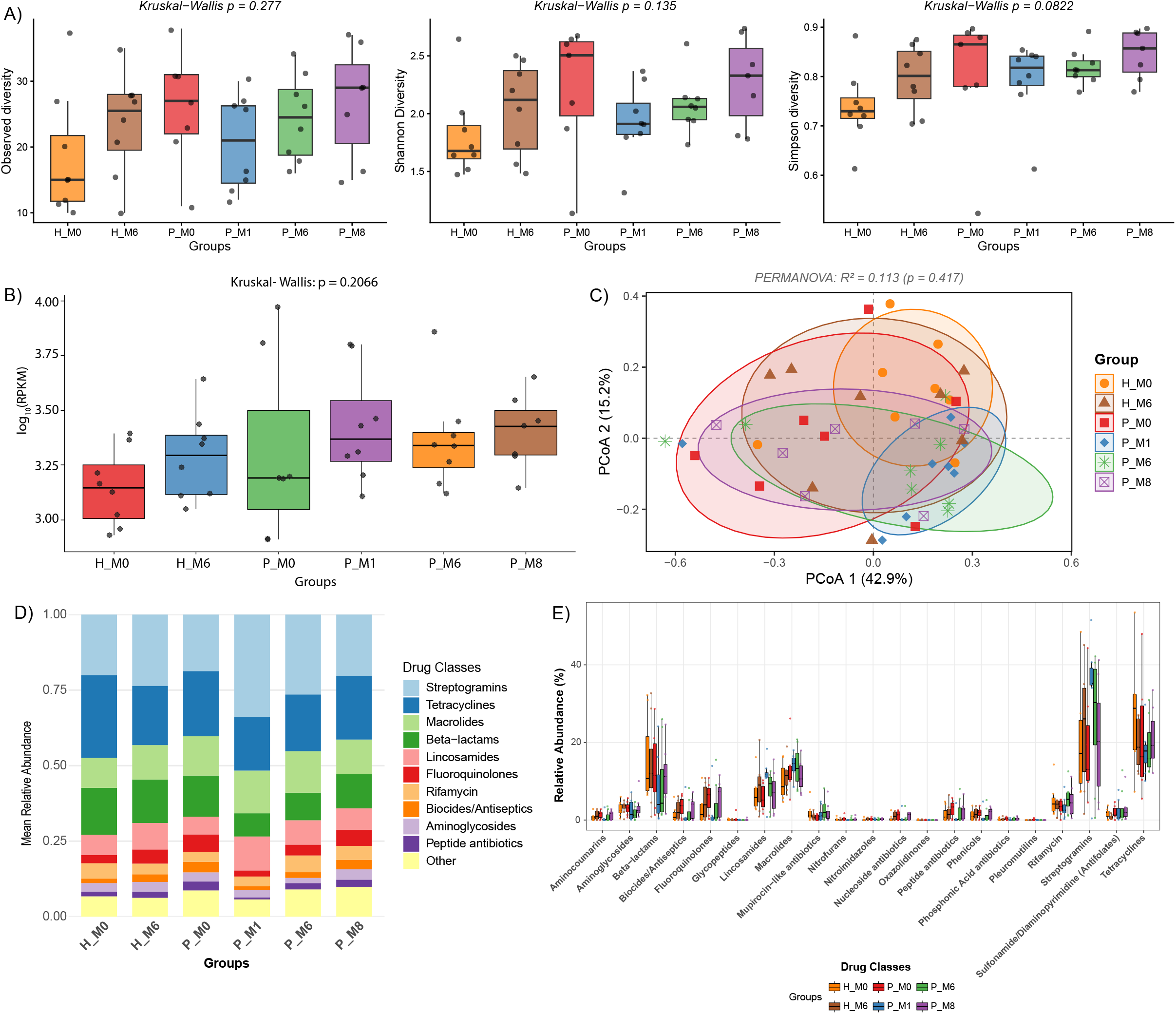
Dynamics of Gut Antimicrobial Resistome Diversity and Drug Class Abundance. A) Alpha Diversity of AMR Genes: Comparison of resistome alpha diversity metrics across study cohorts. Boxplots depict median, IQR, and range with overlaid sample points (global Kruskal-Wallis test: Observed p = 0.277, Shannon p = 0.135, Simpson p = 0.0822). B) Total AMR Gene Abundance: Relative abundance of overall AMR genes quantified as log_10_(RPKM) across study time points (global Kruskal-Wallis test: p = 0.2066). C) Beta Diversity of Resistome Profiles: PCoA plot based on resistome composition showing overall community similarity across cohorts (PC1 = 42.9%, PC2 = 15.2%; PERMANOVA R^2^ = 0.113, p = 0.417; Dispersion p = 0.8984). D) Mean Relative Abundance of Top 10 Drug Classes: Stacked bar chart illustrating the compositional distribution of the top 10 drug resistance classes across groups, dominated by Streptogramins, Tetracyclines, Macrolides, and Beta-lactams. E) Distribution Across All Resistance Classes: Boxplots showing percentage relative abundance comparisons across specific antimicrobial classes between healthy baseline/follow-up and patient groups over time.

Next, the overall abundance of antimicrobial resistance genes (ARGs), quantified as log_10_-transformed Reads Per Kilobase million (RPKM), also remained comparable across the cohorts (Kruskal-Wallis, p = 0.2066) (Fig. 5B). This suggested a largely stable collective quantitative load. Healthy (H_M0 / H_M6) and untreated patients (P_M0) groups maintained lower, stable resistome burdens. Exploratory pairwise analysis indicated a transient increase following ATT initiation, with ARG abundance escalated at month 1 (P_M1 vs. H_M0: p_unadj = 0.034), declining by month 6, and remaining moderately elevated at month 8 (P_M8 vs. H_M0, p_unadj = 0.029), relative to H_M0. These unadjusted variations (p_adj > 0.05) nominally tracked a transient, treatment-induced inflation of the total intestinal resistance reservoir before stabilizing in the shared household environment.

To investigate overall resistome composition, we performed PCoA based on beta diversity. The first two principal coordinates explained 64.5% of the total community variance (PC1 = 42.9%, PC2 = 15.2%) (Fig. 5C). Extensive overlap among study groups was observed, consistent with the absence of significant differences in global resistome composition (PERMANOVA: R² = 0.113, p = 0.417). Likewise, beta-dispersion analysis indicated comparable within-group variability (Betadisper: p > 0.05, p_adj ∼ 0.918). Post-hoc pairwise comparisons, utilizing stratified paired tests for longitudinal intra-subject monitoring and unpaired tests for cross-cohort comparisons, confirmed a lack of significance across all community centroids (PERMANOVA: p > 0.05, p_adj > 0.8). The most notable nominal variation occurred during early treatment (P_M0 vs. P_M1; R^2^ = 0.105, p_unadj = 0.055, p_adj = 0.820). Overall, these findings indicate that the global structure of the gut resistome remained largely stable throughout treatment. Finally, compositional analysis revealed that resistance profiles were persistently dominated by four major antibiotic classes: Streptogramins, Tetracyclines, Macrolides and Beta-lactams (Fig. 5D), which collectively accounted for over 50% of the total relative abundance across all groups. While the healthy controls (H_M0 vs. H_M6) exhibited a stable distribution of core drug classes over time, the patient cohort showed a distinct, transient compositional shift at month 1 (P_M1). This phase displayed a transient increase in streptogramin resistance genes at month 1, alongside a concomitant suppression of beta-lactam and tetracycline resistance genes. This pattern alteration gradually reverted toward baseline by month 6 (P_M6) and stabilized by month 8 (P_M8) (Figs. 5D-E). Ultimately, these data indicate that while ATT was associated with transient fluctuations in the abundance of specific ARG classes, the overall resistome composition remained relatively resilient during the study period.

### 9) Quantitative Architecture and Diversity Dynamics of the Gut Mobilome

To evaluate the impact of Mtb infection, ATT, and shared domestic environments on the intestinal mobilome, the longitudinal diversity, composition, and abundance of plasmid and viral populations across the study cohorts were analysed.

#### Analysing the Gut Plasmidome

Global plasmid alpha diversity did not differ significantly across the study groups (Kruskal-Wallis; Obs.: p = 0.111, Shan.: p = 0.0738, Simp.: p = 0.103) (Fig. 6A). Nevertheless, exploratory pairwise analyses suggested localized differences. Healthy household contacts maintained relatively stable plasmid diversity over six months, whereas patients exhibited greater inter-individual variability and a nominal reduction in alpha diversity during early treatment, most prominently at month 1 (H_M0 vs. P_M0: Obs. p_unadj = 0.02, Shan. p_unadj = 0.009, Simp. p_unadj = 0.007; H_M0 vs. P_M1: Obs. p_unadj = 0.010, Shan. p_unadj = 0.008, Simp. p_unadj = 0.021). Plasmid diversity gradually increased during follow-up (P_M6 to P_M8), approaching baseline levels. However, none of these pairwise differences remained significant following FDR correction (p_adj > 0.05), they point to a transient, treatment-induced compression of specific plasmid-carrying lineages before recovery.

**Figure 6.**
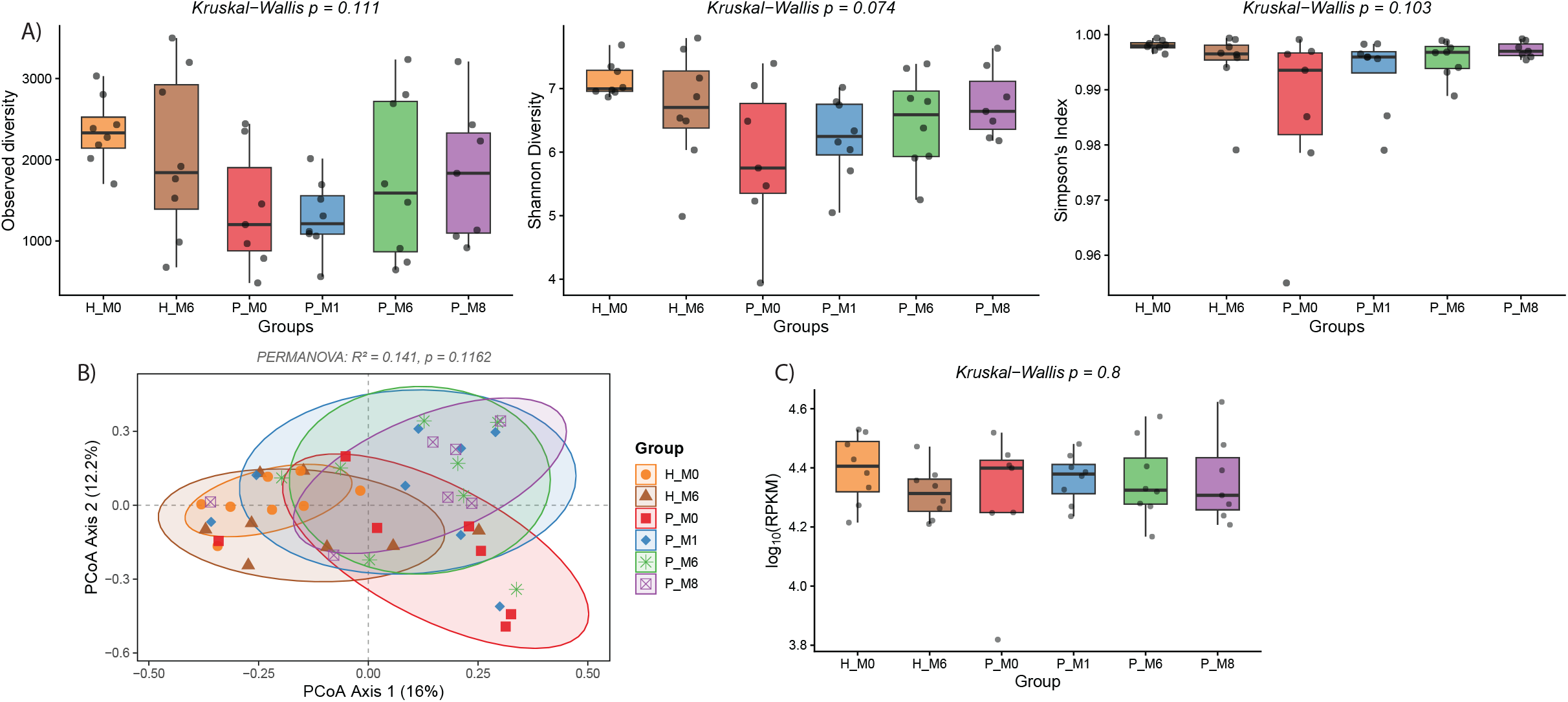
Diversity and Abundance Dynamics of Plasmid-Associated Mobile Genetic Elements (MGEs). A) Alpha Diversity of Plasmid MGEs: Comparison of plasmid MGE alpha diversity metrics across study groups. Boxplots display the median, interquartile range (IQR), and full range with individual data points overlaid for Observed diversity (p = 0.111), Shannon diversity (p = 0.074), and Simpson’s Index (p = 0.103; global Kruskal-Wallis tests). B) Beta Diversity of Plasmid Composition: PCoA plot based on plasmid MGE profiles across healthy and patient cohorts (PCoA 1 = 16%, PCoA 2 = 12.2% variance explained, PERMANOVA R^2^ = 0.141, p = 0.1162). C) Total Plasmid MGE Abundance: Boxplot illustrating overall abundance of plasmid-associated MGEs quantified as log_10_(RPKM) across cohorts over time (global Kruskal-Wallis test: p = 0.8).

To assess plasmid beta diversity, PCoA was performed, capturing a combined 28.2% of total community variance (PCoA1 = 16.0%, PCoA2 = 12.2%) (Fig. 6B). While the global PERMANOVA was non-significant (R^2^ = 0.141, p = 0.1162), pairwise comparisons reveal distinct plasmid community dynamics among study groups. At baseline, patient plasmid composition differed significantly from healthy controls (P_M0 vs H_M0: p = 0.0134, p_adj = 0.0430), although with significant dispersion heterogeneity (p = 0.0001, p_adj = 0.0015). Over time, plasmid composition shifted from baseline to month 6 (P_M0 vs P_M6: p = 0.0156, p_adj = 0.0430; dispersion p = 0.3314), ultimately establishing a distinct plasmid profile compared to month 6 healthy controls (P_M6 vs H_M6: p = 0.0163, p_adj = 0.0430) under homogeneous dispersion conditions (p = 0.3918). Furthermore, longitudinal patient plasmid composition remained significantly distinct from baseline controls (H_M0). This was evident at month 1 (P_M1 vs H_M0: p = 0.0172, p_adj = 0.0430), month 6 (P_M6 vs H_M0: p = 0.0013, p_adj = 0.0195), and month 8 (P_M8 vs H_M0: p = 0.0113, p_adj = 0.0430). While the differences at month 1 and month 6 are accompanied by significant group variance heterogeneity (dispersion p_adj ≤ 0.025), the divergence at month 8 persists as a true structural shift under stable, homogeneous dispersion (p_adj = 0.1028).

In contrast, total plasmid abundance, quantified as log_10_-transformed RPKM, remained stable across all groups (Kruskal–Wallis, p = 0.816), and no pairwise comparison reached statistical significance (Fig. 6C). These findings indicate that although plasmid community composition changed during treatment, the overall plasmid load remained conserved.

#### Analysing the Gut Virome

The alpha diversity of viral mobile genetic elements (MGE-viruses) also remained globally comparable across groups (Kruskal-Wallis: Obs. p = 0.07, Shan. p = 0.0813, Simp. p = 0.126) (Fig. 7A). Exploratory pairwise comparisons revealed a reduction in baseline viral alpha diversity in treatment-naive patients relative to controls (Obs. H_M0 vs P_M0: p_unadj = 0.0231; H_M6 vs P_M0: p_unadj = 0.0076; Shan. H_M0 vs P_M0: p_unadj = 0.0105; Simp. H_M0 vs P_M0: p_unadj = 0.0119). Within the patient cohort, the viral alphadiversity increased during treatment (P_M0 vs P_M6: Obs. p_unadj = 0.0399; Shan. p_unadj = 0.0348; Simp. p_unadj = 0.0405) and by months 6 and 8, becoming comparable to healthy controls.

**Figure 7.**
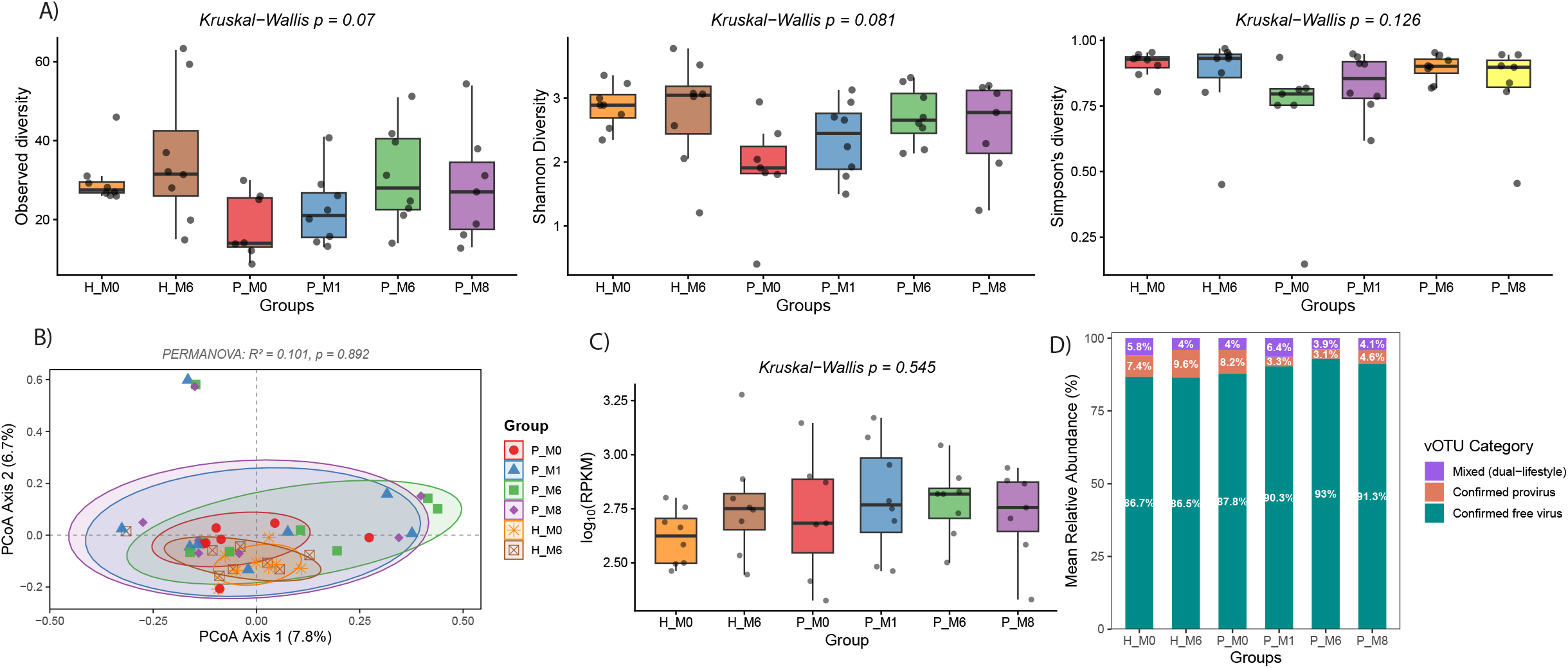
Profiling Gut Viral Mobile Genetic Elements (MGEs) Diversity, Abundance, and Lifestyle Categories. A) Alpha Diversity of Viral MGEs: Comparisons of viral MGE alpha diversity metrics across study groups. Boxplots display the median, interquartile range (IQR), and range with individual sample data points overlaid for Observed diversity (p = 0.07), Shannon diversity (p = 0.0813), and Simpson’s Index (p = 0.126; global Kruskal-Wallis tests). B) Beta Diversity of Viral Composition: PCoA plot based on viral MGE profiles showing community ordination across healthy and patient cohorts over time (PCoA 1 = 7.8%, PCoA 2 = 6.7% variance explained, PERMANOVA R^2^ = 0.101, p = 0.892). C) Total Viral MGE Abundance: Boxplot depicting overall abundance of viral-associated MGEs quantified as log_10_(RPKM) across study groups (global Kruskal-Wallis test: p = 0.545). D) Viral OTU (vOTU) Lifestyle Distribution: Stacked bar chart showing the mean relative abundance percentage of vOTU lifestyle categories across study groups, categorized into confirmed free virus (blue), confirmed provirus (orange), and mixed (dual-lifestyle) (purple).

PCoA showed substantial overlap among viral communities, with the first two axes explaining 14.5% of the total variance (PCoA1 = 7.8%, PCoA2 = 6.7%) (Fig. 7B). Consistent with this pattern, global PERMANOVA did not identify significant differences among groups (R² = 0.101, p = 0.861). Pairwise comparisons detected nominal compositional differences between H_M0 and patients at months 1 (p_unadj = 0.0241), 6 (p_unadj = 0.0412), and 8 (p_ unadj = 0.0038), although the divergence at month 1 is partially confounded by significant dispersion heterogeneity (p_unadj = 0.0457), the compositional differences at months 6 and 8 occured under homogeneous dispersion (p_unadj ≥ 0.1528). Also, no significant longitudinal differences were observed within the patient cohort or between patients and time-matched healthy cohort (H_M6).

Total viral abundance also remained stable throughout the study (Kruskal–Wallis, p = 0.545), with no significant pairwise differences in log_10_-transformed RPKM (Fig. 7C). Together, these results indicate that the overall quantity of viral-MGEs was maintained despite disease progression and treatment. To profile the shifting dynamics of the viral community, viral operational taxonomic units (vOTUs) were classified as free viruses, proviruses, or mixed (dual-lifestyle) populations. Free viruses consistently dominated the gut virome across all groups, representing 86.46–93.01% of total viral abundance (Fig. 7D). In contrast, proviruses remained relatively stable in healthy controls (H_M0, 7.42%; H_M6, 9.56%) but declined in patients following ATT initiation, decreasing from 8.20% at P_M0 to 3.33% at P_M1 and 3.11% at P_M6 before partially recovering to 4.63% at P_M8. Mixed viral populations represented a minor but stable fraction of the virome throughout the study (3.88–6.38%). These findings indicate that ATT was associated with transient alterations in viral lifestyle composition, whereas the overall abundance of the gut virome remained stable.

### 10) Longitudinal Profiling of Bacterial–AMR Co-occurrence Networks

To evaluate potential species-ARG associations, Spearman’s rank correlation analysis was performed across all study groups. The resulting co-occurrence networks revealed distinct baseline profiles, temporal trajectories, and clear taxonomic segregation between pathogens and commensals. The healthy baseline (H_M0) exhibited a sparse, modular network with limited pairings, notably *Klebsiella* species with *pmr* PETN transferase, *Bifidobacterium* species with resistant *ileS* synthetase, and specific taxa (*B. ovatus*, *D. formicigenerans*, *E. coli*, *M. gnavus*) with *sul* (Fig. 8A). In contrast, the patient baseline (P_M0) displayed a denser resistome, marked by broad multidrug-resistant (MDR) correlation blocks in *E. coli* spanning multiple efflux systems (SMR, MFS, RND, ABC), porin alterations, *fosfomycin thiol transferase*, *LNU nucleotidyltransferase*, and *MPH phosphotransferase*, alongside secondary clusters involving *S. salivarius*, *V. atypica*, and *R. ilealis* (Fig. 8B). Over time, both cohorts demonstrated network expansion along distinct trajectories. In healthy cohort (H_M6), the network broadened through a cluster of Bacillota (*Streptococcus* species) co-associating with RND/MFS efflux pumps, *kdpDE*, and porin permeability features (Fig. 8C). In the patient cohorts undergoing treatment (P_M1 and P_M6), associations expanded to additional opportunistic Pseudomonadota, including *Pseudomonas* species, *A. xylosoxidans*, and *E. cloacae*, linked to *qnr* quinolone resistance, *cfxA* beta-lactamase, and *mph* phosphotransferase (Fig. 8D-E). By month 8 (P_M8), high-density MDR clusters re-concentrated within core Gram-negative pathogens (*E. coli* and *K. pneumoniae*), whereas commensal taxa across both cohorts (*Bacteroides*, *Veillonella*, *Faecalibacterium*, *Phocaeicola*, *Dorea*, *Blautia*, and *Bifidobacterium*) consistently maintained narrow, low-density linkages restricted to single determinants such as *sul*, *kdpDE*, *ileS synthetase*, *ant(6)*, or macrolide esterases (Fig. 8F).

**Figure 8.**
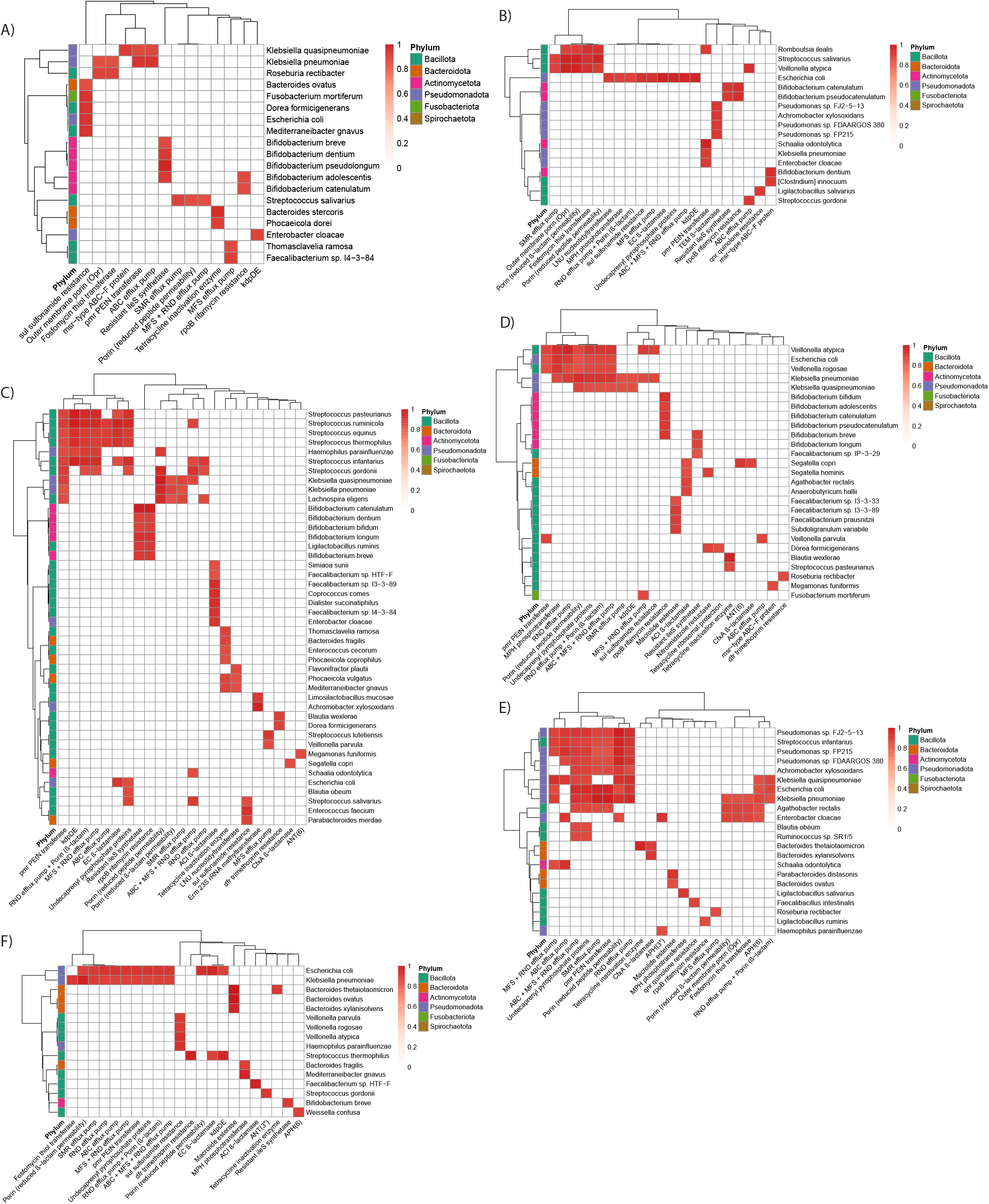
Co-occurrence heatmaps of bacterial species and antimicrobial resistance (AMR) mechanisms across study groups. (A-F) Heatmaps display Spearman’s rank correlation networks for HM0, PM0, HM6, PM1, PM6, and PM8 cohorts. Rows represent bacterial taxa color-coded by phyla, and columns denote specific AMR mechanisms. Correlation strength is indicated by the red color gradient scale (0 to 1). Both rows and columns are hierarchically clustered based on similarity in association profiles.

## DISCUSSION

Tuberculosis (TB) is increasingly recognized as a systemic disease that extends beyond the lungs through bidirectional gut-lung interactions, whereby pulmonary infection perturbs the intestinal microbiome and the gut microbiota, in turn, influences systemic immunity and susceptibility to respiratory disease^26,27^. While previous studies have primarily focused on cross-sectional taxonomic alterations, the longitudinal dynamics of microbial, functional and metabolic recovery during anti-tuberculosis therapy (ATT) remain poorly understood. By integrating longitudinal shotgun metagenomics with functional, metabolomic and mobilome analyses, our study provides a systems-level view of gut ecosystem remodeling during TB and treatment. We demonstrate that active TB is associated with marked perturbations in intestinal microbial composition, function and metabolism, accompanied by alterations in mobile genetic elements (MGEs). Although ATT progressively restored microbial diversity and metabolic homeostasis, the gut ecosystem remained distinct from healthy controls after treatment, suggesting that ecological recovery lags behind microbiological cure.

A major strength of this study is the inclusion of healthy household contacts (HHCs) rather than unrelated healthy controls, minimizing confounding from shared environmental, dietary and socioeconomic factors while accounting for continuous Mtb exposure. Consequently, the observed microbiome alterations are more likely to reflect disease-associated changes than environmental variation. Despite prolonged exposure to index TB cases, HHCs demonstrated relative stability of gut microbial, functional and metabolic profiles over time, with only modest longitudinal variations observed. Although our study was not designed to identify protective microbial signatures, these findings are consistent with the hypothesis that preservation of gut homeostasis may contribute to resilience against progression to active TB through the gut-lung axis^3,28,29^. Future longitudinal studies comparing exposed HHCs who progress to active disease with those who remain healthy may help identify microbiome-derived biomarkers of TB susceptibility or natural resistance.

Active pulmonary TB patients had significantly lower BMI than HHCs, reflecting disease-associated wasting and chronic inflammation^30,31^. ATT was accompanied by gradual BMI recovery and significant declines in total WBCs, neutrophil, and monocyte counts, indicating resolution of systemic inflammation with treatment.

A principal finding of this study is that active TB is associated with intestinal dysbiosis, characterized by reduced microbial diversity, altered community structure and increased inter-individual variability. Treatment-naive patients exhibited significantly lower alpha diversity, distinct microbial community composition and greater beta dispersion than healthy household contacts, indicating disruption of gut microbial homeostasis. These findings are consistent with previous studies reporting reduced microbial diversity in pulmonary TB^3,28,32^. The increased community dispersion likely reflects heterogeneity in disease severity, inflammatory status and host responses, supporting the concept that pulmonary Mtb infection induces systemic alterations in gut microbial ecology through the gut-lung axis.

Taxonomic profiling identified candidate alterations in the gut microbiota during active TB, characterized by depletion of beneficial commensals, including *Faecalibacterium prausnitzii*, *Bifidobacterium* spp., *Blautia* spp., *Collinsella aerofaciens*, *Parabacteroides distasonis*, *Bacteroides ovatus* and *Limosilactobacillus mucosae*. These taxa contribute to complex carbohydrate fermentation, SCFA production, bile acid metabolism, epithelial barrier maintenance and immune homeostasis^25,33^. Their depletion therefore suggests impaired microbial fermentation and disruption of host-microbe metabolic interactions. Conversely, treatment-naive patients exhibited enrichment of opportunistic and inflammation-associated taxa, including *Enterococcus faecium*, *Escherichia coli*, *Streptococcus* spp., *Bacteroides fragilis*, *Bacteroides stercoris*, *Thomasclavelia ramose* and *Megamonas funiformis*. Expansion of these organisms has been linked to inflammatory intestinal environments characterized by altered oxygen availability and increased host-derived nutrients^34,35^. Together, these findings indicate a shift from a commensal-rich to an inflammation-adapted gut ecosystem, consistent with the accompanying metabolomic alterations observed in our study.

Longitudinal analysis showed that ATT progressively restored microbial diversity and reduced ecological instability, although recovery remained incomplete after treatment. Alpha diversity increased over time, and beta diversity shifted towards healthy household contacts without complete convergence, indicating persistent alterations in community composition. This recovery was accompanied by the re-emergence of beneficial SCFA-producing taxa, including *Faecalibacterium prausnitzii*, *Bifidobacterium longum*, *Anaerobutyricum hallii*, *Blautia* spp. and *Parabacteroides distasonis*, consistent with restoration of fermentative metabolism and gut homeostasis. However, the persistence of several opportunistic taxa suggests that microbiological cure does not necessarily coincide with complete ecological recovery. Similar delayed restoration of the gut microbiome following prolonged antibiotic exposure has been reported in both TB and non-TB settings^28,36^.

The persistent separation between treated patients and HHCs is notable, as HHCs represent an environmentally matched healthy reference state. These findings suggest that ATT and disease-associated perturbations may induce prolonged alterations in gut microbial ecology that persist beyond microbiological cure. Incomplete recovery of SCFA-producing taxa and microbial metabolic functions may contribute to sustained changes in host-microbe interactions and immune-metabolic homeostasis. Thus, restoration of gut ecosystem function may represent an important component of post-TB recovery beyond pathogen clearance.

Beyond taxonomic restructuring, functional metagenomic analysis revealed substantial alterations in the metabolic potential of the gut microbiome during active TB. Treatment-naive patients showed depletion of pathways involved in carbohydrate utilization, amino acid biosynthesis, nucleotide metabolism, vitamin biosynthesis and energy metabolism, accompanied by enrichment of stress adaptation and alternative metabolic processes. These findings indicate that TB-associated dysbiosis extends beyond compositional changes to affect microbial functional capacity. Similar functional shifts have been reported in chronic inflammatory conditions, where loss of metabolically versatile anaerobic commensals is associated with reduced biosynthetic potential and altered microbiota-derived metabolite production^37,38^. Notably, the depletion of several functional pathways coincided with the decline of corresponding microbial taxa, suggesting coordinated taxonomic and functional remodeling of the gut ecosystem during active disease.

Following ATT initiation, restoration of microbial functional pathways paralleled the recovery of beneficial commensals. Pathways involved in central carbon metabolism, amino acid biosynthesis, glycan degradation, cofactor and vitamin biosynthesis gradually increased during treatment, indicating progressive recovery of microbial metabolic capacity. However, several pathways remained distinct from HHCs after treatment completion, suggesting that functional recovery lagged behind bacterial clearance. Given the functional redundancy of the gut microbiome, different microbial communities may maintain similar metabolic functions; however, prolonged depletion of functionally important taxa may compromise ecosystem resilience and metabolic stability^37,39^. These findings highlight that recovery following ATT should be evaluated beyond microbial diversity and include restoration of microbial functional potential.

Fecal metabolomic profiling further supported extensive metabolic remodeling during TB progression and treatment. Active disease was associated with alterations in metabolites involved in amino acid metabolism, lipid metabolism, oxidative stress, bile acid transformation and host-microbiome co-metabolism, followed by gradual normalization during therapy. The concordant recovery of microbial functions and metabolite profiles suggests that taxonomic restructuring translated into biologically relevant metabolic changes. As microbial metabolites influence host immunity, epithelial integrity and systemic metabolism, these findings demonstrate that TB-associated gut alterations extend beyond microbial composition to functional and metabolic consequences^40,41^.

Alterations in amino acid metabolism, bile acid metabolism and oxidative stress-related metabolites were particularly relevant to TB-associated metabolic perturbation. These pathways regulate microbial growth, immune signalling, epithelial integrity and host-microbial metabolic interactions. Although causal relationships cannot be established, the coordinated recovery of microbial taxa, functional pathways and metabolite profiles suggests interconnected restoration of microbial and host metabolic homeostasis.

Integration of shotgun metagenomics and metabolomics revealed coordinated associations between bacterial taxa and metabolites, providing insights into functional interactions within the gut ecosystem. Beneficial commensals showed associations with metabolites linked to amino acid and lipid metabolism and intestinal homeostasis, whereas opportunistic taxa displayed contrasting interaction patterns. Although these correlations do not establish causality, they provide potential mechanistic links between microbial restructuring and metabolic alterations during TB. The systems-level integration of multiple omics layers therefore provides a more comprehensive understanding of host-microbiome interactions and may enable identification of biologically relevant signatures of disease activity and treatment response^38,42^. Future studies incorporating host immune parameters with microbiome and metabolome datasets will further clarify these interactions.

Despite extensive bacterial community restructuring, the gut resistome exhibited greater stability during ATT. Overall AMR gene diversity remained relatively unchanged, although specific resistance determinants showed transient fluctuations. These findings suggest that the gut resistome possesses considerable ecological resilience, potentially maintained through persistence of resistance genes within resident microbial populations and horizontal gene transfer (HGT) networks. Nevertheless, transient enrichment of selected resistance classes indicates that prolonged antimicrobial exposure can modulate specific AMR reservoirs without broadly disrupting resistome architecture, consistent with previous antibiotic exposure studies^43,44^.

Plasmid and viral communities also demonstrated greater stability than bacterial composition during ATT. While modest changes in plasmid and virome composition occurred during treatment, overall plasmid abundance remained relatively stable and viral diversity progressively recovered. These findings suggest that MGEs may be more resilient to antimicrobial perturbation than their bacterial hosts, potentially maintaining HGT capacity despite microbiome restructuring. Given their role in AMR dissemination and bacterial adaptation, further functional characterization of these elements will be important for understanding microbial resilience during prolonged antimicrobial therapy.

Bacterial-AMR co-occurrence analysis revealed a strong taxonomic bifurcation, where multidrug resistance (MDR) mechanisms (including multi-class efflux pumps and porin alterations) concentrated within opportunistic Pseudomonadota (*E. coli*, *Klebsiella* spp.), while gut commensals maintained narrow, single-gene associations. Furthermore, longitudinal expansion across timepoints highlighted dynamic resistome restructuring, suggesting that specific pathobionts serve as primary reservoirs for complex MDR profiles in the gut ecosystem.

This study provides a comprehensive longitudinal characterization of gut ecosystem remodeling during pulmonary TB and ATT. The longitudinal design enabled assessment of disease-associated dysbiosis, treatment-associated recovery and post-treatment ecological persistence. Inclusion of HHCs as environmentally matched controls reduced confounding from geography, diet and lifestyle, allowing better attribution of observed alterations to TB-associated processes. Integration of shotgun metagenomics, functional profiling, metabolomics, microbiome-metabolome interactomics, resistome, plasmidome, virome and bacterial-AMR network analyses provides a multidimensional view of gut ecosystem dynamics. Additionally, longitudinal dietary assessments and hematological parameters enabled interpretation of microbiome changes within the broader clinical and nutritional context of TB treatment.

Despite these strengths, several limitations should be acknowledged. Although the longitudinal design enabled assessment of temporal microbial changes, the modest sample size and single-center recruitment may limit generalizability across diverse populations with different genetic, dietary, environmental and epidemiological backgrounds. While dietary intake and routine hematological parameters were evaluated, comprehensive host immune profiling was not performed. Integration of cytokine profiles, immune cell phenotyping and inflammatory markers with microbiome and metabolomic data will provide deeper mechanistic insights into host-microbiome interactions during TB and treatment. Also, loss of SCFA-producing bacteria may affect intestinal metabolism, however, SCFAs were not quantified in this study. Furthermore, the observational nature of this study precludes causal inference between microbial alterations, metabolic changes and disease progression. Future studies incorporating experimental validation and extended post-treatment follow-up will be essential to determine the long-term clinical relevance of persistent gut ecosystem alterations.

Collectively, our findings support the view of pulmonary TB as a systemic disease associated with extensive remodeling of the intestinal ecosystem. The coordinated alterations in microbial composition, function, metabolism and genetic elements highlight the interconnected nature of host-microbiome interactions during disease and treatment. Persistence of microbial and metabolic differences after completion of ATT suggests that ecological recovery extends beyond microbiological cure, emphasizing the importance of considering restoration of gut homeostasis as a component of post-TB recovery.

## CONCLUSION

In conclusion, our multi-omic study demonstrates that active pulmonary TB induces coordinated alterations in gut microbial composition, function, metabolism, and genetic elements, with only partial recovery following ATT despite microbiological cure. While healthy household contacts exposed to similar environments maintained relative gut ecosystem stability over time, treated patients exhibited persistent ecological differences, indicating that microbiome recovery extends beyond Mtb clearance. These findings highlight the importance of considering restoration of gut ecosystem function as an integral component of post-TB recovery and provide a foundation for future microbiome-directed interventions.

## Data Availability

The metagenomic sequencing data generated in this study have been submitted to the NCBI Sequence Read Archive (SRA) under BioProject accession number BioProject ID PRJNA1531317 and will be released upon publication. Processed datasets supporting the findings of this study are available from the corresponding author upon reasonable request.

## Credit Author Statement

AP, SS, SB, BD & GA conceptualized and designed the study. AP, JS, SKY, SKG, PJ & DT performed experiments and data curation. AP, SKY & BPG performed formal analysis and visualization. Patient recruitment, clinical evaluation, and sample collection was done by SS & SB. GA, YK, BD & DM provided resources. AP analysed the data and wrote the paper. AP, GA, YK, BD & DM reviewed, edited and finalized the article.

## Ethical approval

The study protocol was approved by the ethics committees of BRIC-National Institute of Immunology, New Delhi and Jawaharlal Institute of Postgraduate Medical Education & Research, Puducherry. Participants received information about the purpose of the study and their right to confidentiality. Written informed consent was obtained from each participant.

## Data Availability

The whole-genome shotgun metagenomic data generated in this study has been deposited in the National Center for Biotechnology Information (https://www.ncbi.nlm.nih.gov/), under **BioProject ID PRJNA1531317**.

## Appendix A. Supporting information

Supplementary data associated with this article will be made available upon acceptance of the manuscript.

## Acknowledgements

This study received financial support from Department of Science & Technology (DST), Innovation in Science Pursuit for Inspired Research (INSPIRE)-Faculty Fellowship (Grant Number DST/INSPIRE/04/2020/001246), Govt. of India (GOI). AP received support from DST-INSPIRE Faculty Fellowship, GOI. AP acknowledges BRIC-National Institute of Immunology, for providing space, and research facilities. We acknowledge the Bioinformatics Centre, BRIC-NII for providing access to the computational facility. Authors are grateful to Mr. Khem Singh Negi for technical help.

## Supplementary Figures

**Suppl. Figure S1.**
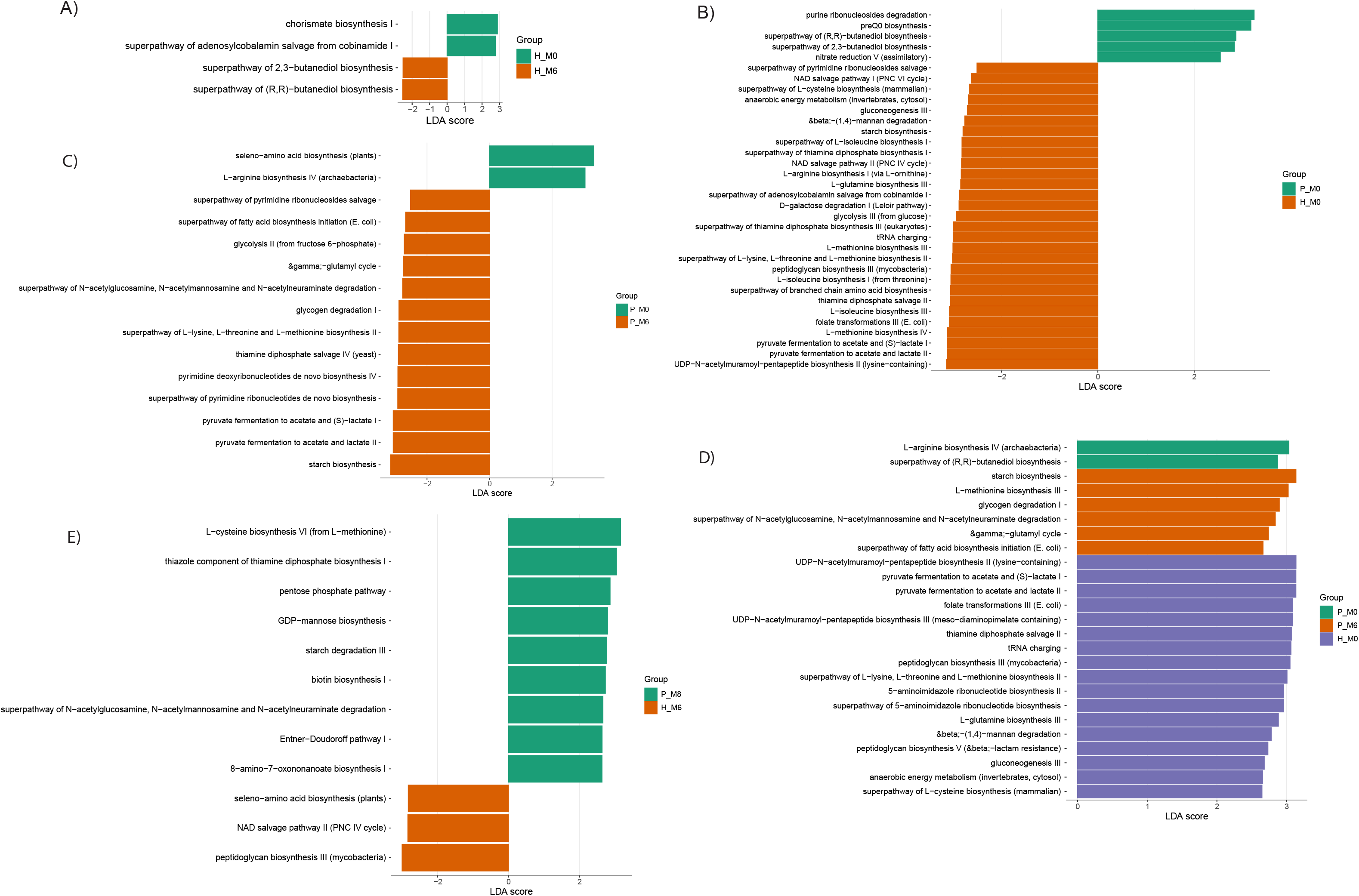
Identification of exploratory functional features associated with tuberculosis and anti-tuberculosis treatment by Linear Discriminant Analysis Effect Size (LEfSe) analysis. Linear Discriminant Analysis (LDA) effect size plots highlighting enriched functional features (pathway levels) across longitudinal baseline and follow-up comparisons between healthy controls (H) and patients (P). Exploratory pathways (LDA score > 2.5, p < 0.05) are highlighted according to group assignment. (A) Longitudinal baseline vs. follow-up in healthy controls (H_M0 vs. H_M6). (B) Baseline profiles depicting dysbiotic shifts between healthy controls (H_M0) and treatment-naive patients (P_M0). (C) Treatment-driven taxonomic and functional shifts (P_M0 vs. P_M6). (D) Multigroup comparison (H_M0 vs. P_M0 vs. P_M6) mapping exploratory profiles across baseline healthy controls, baseline patients, and post-treatment patients. (E) Endpoint Comparison (H_M6 vs. P_M8): Plot identifying persistent variations between healthy follow-up controls (H_M6) and post-treatment follow-up patients (P_M8).

**Suppl. Figure S2.**
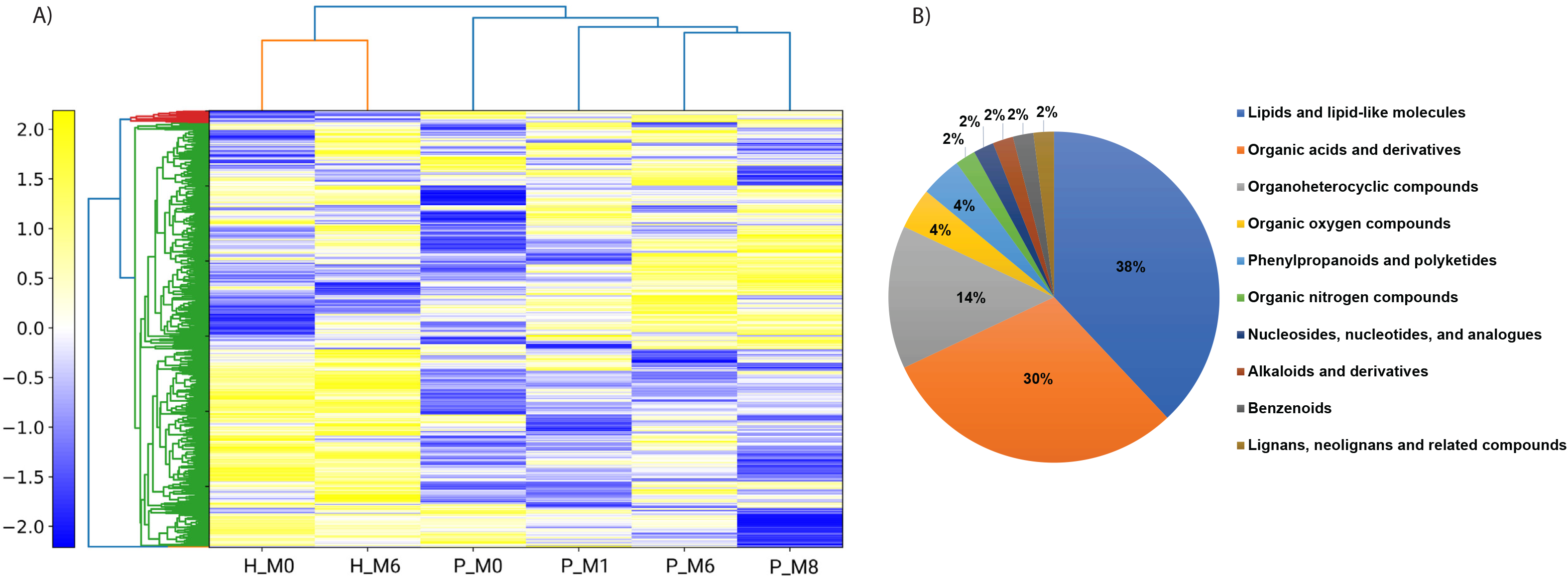
Untargeted Stool Metabolomics Profiling and Differential Exploratory Features Classification. A) Hierarchical Clustering of Global Metabolome: Unsupervised hierarchical clustering heatmap showing overall metabolic signatures across study groups. Dendrograms illustrate global similarity relationships across time points and cohorts (color gradient: Z-score normalized relative abundance; yellow = high, blue = low). B) Chemical Superclass Annotation: Pie chart showing the chemical distribution of identified differential metabolites. Predominant classes include Lipids and lipid-like molecules (38%), Organic acids and derivatives (30%), and Organoheterocyclic compounds (14%).

